## Supplementary material for "Genome-wide association study identifies susceptibility loci for acute myeloid leukemia": Lin 2021 Supplementary Tables

Lin *et al*

**Supplementary Table 1 –Acute myeloid leukemia (AML) case and control characteristics**

N, number; GWAS, genome wide association study; APL, acute promyelocytic leukemia; CBF, core binding factor (CBF AML is defined as t(8;21) or inv(16)); Complex karyotype is defined as 3 or more independent alterations; Monosomal karyotype is defined as the presence of 2 or more autosomal monosomies or a single autosomal monosomy with at least one structural abnormality. For some cases age and/or gender was not declared or not known. Some cytogenetic sub-groups are not mutually exclusive.

|  | **GWAS1** | | **GWAS2** | | **GWAS3** | | **GWAS4** | |
| --- | --- | --- | --- | --- | --- | --- | --- | --- |
|  | **Cases** | **Controls** | **Cases** | **Controls** | **Cases** | **Controls** | **Cases** | **Controls** |
| **Total, N** | 1119 | 2671 | 931 | 2477 | 991 | 1612 | 977 | 3728 |
| **Age, years** |  |  |  |  |  |  |  |  |
| Median (range) | 54 (13-91) | 45 (45-45) | 49 (<1-95) | 45 (15-65) | 60 (12-93) | 62 (32-81) | 63 (<1-93) | 57 (40-70) |
| **Sex, N (%)** |  |  |  |  |  |  |  |  |
| Males | 605 (54) | 1377 (50) | 486 (52) | 1225 (49) | 500 (50) | 785 (49) | 511 (52) | 1746 (47) |
| Females | 514 (46) | 1294 (50) | 444 (48) | 1252 (51) | 478 (48) | 827 (51) | 466 (48) | 1982 (53) |
| Not known | 0 (0) | 0 (0) | 1 (0) | 0 (0) | 13 (2) | 0 (0) | 0 (0) | 0 (0) |
| **Cytogenetics, N (%)** |  |  |  |  |  |  |  |  |
| Successful cytogenetics | 879 (100) | - | 654 (100) | - | 666 (100) | - | 718 (100) | - |
| Normal cytogenetics | 359 (41) | - | 177 (27) | - | 286 (43) | - | 465 (65) | - |
| Abnormal cytogenetics | 520 (59) | - | 477 (73) | - | 380 (57) | - | 253 (35) | - |
| APL t(15.17) | 131 (15) | - | 380 (58) | - | 108 (16) | - | 38 (5) | - |
| CBF AML | 72 (8) | - | 50 (8) | - | 219 (33) | - | 27 (4) | - |
| del5.del7 | 65 (7) | - | 58 (9) | - | 68 (10) | - | 57 (8) | - |
| Complex (3 or more) | 69 (8) | - | 61 (9) | - | 89 (13) | - | 82 (11) | - |
| Any translocation | 241 (27) | - | 459 (70) | - | 253 (38) | - | 105 (15) | - |
| Any trisomy | 106 (12) | - | 70 (11) | - | 55 (8) | - | 104 (14) | - |
| Any monosomy | 54 (6) | - | 56 (9) | - | 67 (10) | - | 62 (9) | - |
| Monosomal karyotype | 20 (2) | - | 34 (5) | - | 54 (8) | - | 46 (6) | - |

**Supplementary Table 2 – SNPs showing evidence of an association with risk of acute myeloid leukemia (AML) at *P* ≤ 10^-6^.**

Results are based on meta-analysis of GWAS1, GWAS2 and GWAS3. SNP, single nucleotide polymorphism; Chr, chromosome; OR, odds ratio; Q, Cochran’s Q statistic; I^2^, heterogeneity index I^2^. ^a^hg19 coordinates

| SNP | **AML karotype** | **Chr** | **Position^a^** | **Chr band** | **Allele 1** | **Allele 2** | **Meta *P* value (fixed effect)** | **Meta P (random effect)** | **Meta OR (fixed effect)** | **Meta OR (random effect)** | **Q** | **I^2^** |
| --- | --- | --- | --- | --- | --- | --- | --- | --- | --- | --- | --- | --- |
| rs4674579 | All AML | 2 | 222167434 | 2q36.1 | C | T | 7.00E-07 | 7.00E-07 | 0.8063 | 0.8063 | 0.63 | 0 |
| rs2621279 | All AML | 3 | 158940084 | 3q25.32 | A | G | 4.63E-07 | 4.63E-07 | 0.815 | 0.815 | 0.7933 | 0 |
| rs13164987 | All AML | 5 | 168247045 | 5q35.1 | C | T | 4.79E-07 | 4.79E-07 | 0.7885 | 0.7885 | 0.6976 | 0 |
| rs13183143 | All AML | 5 | 168247046 | 5q35.1 | A | G | 4.37E-07 | 4.37E-07 | 0.7876 | 0.7876 | 0.6849 | 0 |
| rs11481 | All AML | 11 | 67820335 | 11q13.2 | A | T | 8.04E-07 | 8.04E-07 | 1.2044 | 1.2044 | 0.4994 | 0 |
| rs10896298 | All AML | 11 | 67931459 | 11q13.2 | T | C | 9.83E-07 | 9.83E-07 | 1.1777 | 1.1777 | 0.4718 | 0 |
| rs4930561 | All AML | 11 | 67931761 | 11q13.2 | A | G | 9.26E-07 | 9.26E-07 | 1.1781 | 1.1781 | 0.468 | 0 |
| rs6056038 | All AML | 20 | 8701468 | 20p12.3 | C | T | 8.90E-07 | 8.90E-07 | 0.762 | 0.762 | 0.9602 | 0 |
| rs6056041 | All AML | 20 | 8702867 | 20p12.3 | C | T | 5.41E-07 | 5.41E-07 | 0.7569 | 0.7569 | 0.9746 | 0 |
| rs6077414 | All AML | 20 | 8704552 | 20p12.3 | C | T | 3.77E-07 | 3.77E-07 | 0.7533 | 0.7533 | 0.9702 | 0 |
| rs6056043 | All AML | 20 | 8705496 | 20p12.3 | C | A | 3.94E-07 | 3.94E-07 | 0.7534 | 0.7534 | 0.9756 | 0 |
| rs75391980 | Normal | 4 | 96948521 | 4q22.3 | C | T | 3.40E-08 | 3.40E-08 | 1.7457 | 1.7457 | 0.4139 | 0 |
| rs9275092 | Normal | 6 | 32648987 | 6p21.32 | T | C | 6.47E-07 | 6.47E-07 | 0.5511 | 0.5511 | 0.566 | 0 |
| rs9275095 | Normal | 6 | 32649088 | 6p21.32 | G | C | 5.48E-07 | 5.48E-07 | 0.5456 | 0.5456 | 0.5911 | 0 |
| rs9275097 | Normal | 6 | 32649126 | 6p21.32 | G | A | 4.48E-07 | 4.48E-07 | 0.5486 | 0.5486 | 0.5575 | 0 |
| rs9275098 | Normal | 6 | 32649161 | 6p21.32 | T | C | 4.88E-07 | 4.88E-07 | 0.553 | 0.553 | 0.5503 | 0 |
| rs9275167 | Normal | 6 | 32653263 | 6p21.32 | G | A | 4.94E-07 | 4.94E-07 | 0.5616 | 0.5616 | 0.6229 | 0 |
| rs9275184 | Normal | 6 | 32654714 | 6p21.32 | C | T | 5.22E-07 | 5.22E-07 | 0.5631 | 0.5631 | 0.6254 | 0 |
| rs9275203 | Normal | 6 | 32656947 | 6p21.32 | A | G | 4.92E-07 | 4.92E-07 | 0.5623 | 0.5623 | 0.6321 | 0 |
| rs9275207 | Normal | 6 | 32657710 | 6p21.32 | G | A | 4.36E-07 | 4.36E-07 | 0.5603 | 0.5603 | 0.6523 | 0 |
| rs9275213 | Normal | 6 | 32658335 | 6p21.32 | C | T | 4.39E-07 | 4.39E-07 | 0.5605 | 0.5605 | 0.6568 | 0 |
| rs9275214 | Normal | 6 | 32658665 | 6p21.32 | A | G | 4.47E-07 | 4.47E-07 | 0.5606 | 0.5606 | 0.6594 | 0 |
| rs9275221 | Normal | 6 | 32659099 | 6p21.32 | C | T | 4.62E-07 | 4.62E-07 | 0.5609 | 0.5609 | 0.6583 | 0 |
| rs9275223 | Normal | 6 | 32659839 | 6p21.32 | A | G | 4.68E-07 | 4.68E-07 | 0.5611 | 0.5611 | 0.6645 | 0 |
| rs9275275 | Normal | 6 | 32662607 | 6p21.32 | A | G | 4.94E-07 | 4.94E-07 | 0.5616 | 0.5616 | 0.6731 | 0 |
| rs9275293 | Normal | 6 | 32663308 | 6p21.32 | C | T | 5.06E-07 | 5.06E-07 | 0.5619 | 0.5619 | 0.6777 | 0 |
| rs9275296 | Normal | 6 | 32663431 | 6p21.32 | C | T | 5.08E-07 | 5.08E-07 | 0.562 | 0.562 | 0.6791 | 0 |
| rs9275300 | Normal | 6 | 32664126 | 6p21.32 | G | A | 4.47E-07 | 4.47E-07 | 0.5598 | 0.5598 | 0.6864 | 0 |
| rs9275307 | Normal | 6 | 32664990 | 6p21.32 | T | A | 5.11E-07 | 5.11E-07 | 0.562 | 0.562 | 0.678 | 0 |
| rs9275308 | Normal | 6 | 32665255 | 6p21.32 | A | T | 5.10E-07 | 5.10E-07 | 0.5616 | 0.5616 | 0.6714 | 0 |
| rs9275310 | Normal | 6 | 32665319 | 6p21.32 | G | T | 4.96E-07 | 4.96E-07 | 0.5614 | 0.5614 | 0.6725 | 0 |
| rs9275311 | Normal | 6 | 32665640 | 6p21.32 | T | G | 4.97E-07 | 4.97E-07 | 0.5615 | 0.5615 | 0.6729 | 0 |
| rs9275313 | Normal | 6 | 32665759 | 6p21.32 | T | G | 4.25E-07 | 4.25E-07 | 0.5594 | 0.5594 | 0.662 | 0 |
| rs9275314 | Normal | 6 | 32665909 | 6p21.32 | A | C | 5.17E-07 | 5.17E-07 | 0.5618 | 0.5618 | 0.6682 | 0 |
| rs9275315 | Normal | 6 | 32665912 | 6p21.32 | A | C | 5.17E-07 | 5.17E-07 | 0.5618 | 0.5618 | 0.6687 | 0 |
| rs9275324 | Normal | 6 | 32666635 | 6p21.32 | T | C | 4.51E-07 | 4.51E-07 | 0.5577 | 0.5577 | 0.6839 | 0 |
| rs9275325 | Normal | 6 | 32666651 | 6p21.32 | T | G | 4.97E-07 | 4.97E-07 | 0.5585 | 0.5585 | 0.6767 | 0 |
| rs9275326 | Normal | 6 | 32666660 | 6p21.32 | T | C | 5.05E-07 | 5.05E-07 | 0.5588 | 0.5588 | 0.6767 | 0 |
| rs9275334 | Normal | 6 | 32667107 | 6p21.32 | C | T | 5.06E-07 | 5.06E-07 | 0.5616 | 0.5616 | 0.6756 | 0 |
| rs9275373 | Normal | 6 | 32668411 | 6p21.32 | A | G | 2.40E-07 | 2.40E-07 | 0.5639 | 0.5639 | 0.4842 | 0 |
| rs9275382 | Normal | 6 | 32668831 | 6p21.32 | C | T | 2.54E-07 | 2.54E-07 | 0.5419 | 0.5419 | 0.8059 | 0 |
| rs9275383 | Normal | 6 | 32668846 | 6p21.32 | T | G | 2.49E-07 | 2.49E-07 | 0.5483 | 0.5483 | 0.7753 | 0 |
| rs9275387 | Normal | 6 | 32669013 | 6p21.32 | C | T | 5.18E-07 | 5.18E-07 | 0.5619 | 0.5619 | 0.6764 | 0 |
| rs9275394 | Normal | 6 | 32669454 | 6p21.32 | C | A | 5.12E-07 | 5.12E-07 | 0.5616 | 0.5616 | 0.6776 | 0 |
| rs9275395 | Normal | 6 | 32669483 | 6p21.32 | G | A | 5.10E-07 | 5.10E-07 | 0.5615 | 0.5615 | 0.6777 | 0 |
| rs9275400 | Normal | 6 | 32669761 | 6p21.32 | G | C | 5.16E-07 | 5.16E-07 | 0.5619 | 0.5619 | 0.6768 | 0 |
| rs9275422 | Normal | 6 | 32670548 | 6p21.32 | A | G | 5.03E-07 | 5.03E-07 | 0.5616 | 0.5616 | 0.6713 | 0 |
| rs9275426 | Normal | 6 | 32670912 | 6p21.32 | C | T | 5.10E-07 | 5.10E-07 | 0.5615 | 0.5615 | 0.6725 | 0 |
| rs9275429 | Normal | 6 | 32671057 | 6p21.32 | A | G | 5.11E-07 | 5.11E-07 | 0.5616 | 0.5616 | 0.6756 | 0 |
| rs9275430 | Normal | 6 | 32671086 | 6p21.32 | T | C | 5.13E-07 | 5.13E-07 | 0.5617 | 0.5617 | 0.674 | 0 |
| rs9275434 | Normal | 6 | 32671247 | 6p21.32 | T | C | 5.13E-07 | 5.13E-07 | 0.5617 | 0.5617 | 0.6709 | 0 |
| rs9275435 | Normal | 6 | 32671332 | 6p21.32 | C | G | 5.12E-07 | 5.12E-07 | 0.5616 | 0.5616 | 0.6704 | 0 |
| rs9275437 | Normal | 6 | 32671412 | 6p21.32 | T | C | 5.13E-07 | 5.13E-07 | 0.5616 | 0.5616 | 0.6703 | 0 |
| rs9275442 | Normal | 6 | 32671755 | 6p21.32 | A | G | 5.09E-07 | 5.09E-07 | 0.5616 | 0.5616 | 0.6708 | 0 |
| rs9275476 | Normal | 6 | 32672624 | 6p21.32 | C | T | 5.40E-07 | 5.40E-07 | 0.5615 | 0.5615 | 0.6702 | 0 |
| rs9275477 | Normal | 6 | 32672641 | 6p21.32 | C | A | 5.74E-07 | 5.74E-07 | 0.5516 | 0.5516 | 0.721 | 0 |
| rs9275486 | Normal | 6 | 32673099 | 6p21.32 | A | T | 5.34E-07 | 5.34E-07 | 0.5621 | 0.5621 | 0.6669 | 0 |
| rs9275490 | Normal | 6 | 32673385 | 6p21.32 | G | C | 5.37E-07 | 5.37E-07 | 0.5622 | 0.5622 | 0.6672 | 0 |
| rs9275495 | Normal | 6 | 32673574 | 6p21.32 | T | A | 5.59E-07 | 5.59E-07 | 0.5626 | 0.5626 | 0.6731 | 0 |
| rs9275503 | Normal | 6 | 32673931 | 6p21.32 | G | A | 6.37E-07 | 6.37E-07 | 0.5566 | 0.5566 | 0.7228 | 0 |
| rs9275530 | Normal | 6 | 32675523 | 6p21.32 | C | G | 5.52E-07 | 5.52E-07 | 0.5627 | 0.5627 | 0.6811 | 0 |
| rs9275532 | Normal | 6 | 32675634 | 6p21.32 | G | C | 5.47E-07 | 5.47E-07 | 0.5625 | 0.5625 | 0.6785 | 0 |
| rs9275541 | Normal | 6 | 32676159 | 6p21.32 | G | C | 5.40E-07 | 5.40E-07 | 0.5623 | 0.5623 | 0.6747 | 0 |
| rs4273728 | Normal | 6 | 32678491 | 6p21.32 | C | T | 4.82E-07 | 4.82E-07 | 0.5606 | 0.5606 | 0.6383 | 0 |
| rs9275583 | Normal | 6 | 32680299 | 6p21.32 | A | G | 4.18E-07 | 4.18E-07 | 0.5587 | 0.5587 | 0.6045 | 0 |
| rs9275592 | Normal | 6 | 32680620 | 6p21.32 | T | G | 4.16E-07 | 4.16E-07 | 0.5586 | 0.5586 | 0.5971 | 0 |
| rs7454108 | Normal | 6 | 32681483 | 6p21.32 | C | T | 3.77E-07 | 3.77E-07 | 0.5575 | 0.5575 | 0.5786 | 0 |
| rs3957146 | Normal | 6 | 32681530 | 6p21.32 | C | T | 3.76E-07 | 3.76E-07 | 0.5575 | 0.5575 | 0.5777 | 0 |
| rs3998159 | Normal | 6 | 32682019 | 6p21.32 | C | A | 5.21E-07 | 5.21E-07 | 0.5625 | 0.5625 | 0.5693 | 0 |
| rs3957148 | Normal | 6 | 32682137 | 6p21.32 | G | A | 3.15E-07 | 3.15E-07 | 0.5546 | 0.5546 | 0.6094 | 0 |
| rs9275599 | Normal | 6 | 32682429 | 6p21.32 | T | C | 4.58E-08 | 4.58E-08 | 0.5071 | 0.5071 | 0.6565 | 0 |
| rs3997854 | Normal | 6 | 32682915 | 6p21.32 | G | T | 4.18E-09 | 2.78E-08 | 0.5204 | 0.5172 | 0.3289 | 10.06 |
| rs3873448 | Normal | 6 | 32683055 | 6p21.32 | T | C | 8.51E-09 | 9.63E-06 | 0.5296 | 0.5117 | 0.1801 | 41.67 |
| rs3873453 | Normal | 6 | 32683382 | 6p21.32 | T | C | 3.95E-07 | 1.15E-05 | 0.5858 | 0.5758 | 0.2628 | 25.17 |
| rs9275607 | Normal | 6 | 32683645 | 6p21.32 | G | A | 1.10E-07 | 1.10E-07 | 0.5883 | 0.5883 | 0.6732 | 0 |
| rs9275610 | Normal | 6 | 32683750 | 6p21.32 | C | T | 7.52E-08 | 7.52E-08 | 0.5838 | 0.5838 | 0.6345 | 0 |
| rs9275611 | Normal | 6 | 32683763 | 6p21.32 | A | G | 2.01E-08 | 1.82E-06 | 0.5405 | 0.5289 | 0.252 | 27.45 |
| rs9275613 | Normal | 6 | 32684105 | 6p21.32 | T | C | 2.07E-08 | 1.47E-06 | 0.5444 | 0.5335 | 0.259 | 25.98 |
| rs9275614 | Normal | 6 | 32684257 | 6p21.32 | G | A | 2.11E-08 | 1.89E-06 | 0.5451 | 0.5334 | 0.2516 | 27.54 |
| rs9275615 | Normal | 6 | 32684272 | 6p21.32 | A | T | 2.13E-08 | 1.81E-06 | 0.5452 | 0.5337 | 0.253 | 27.25 |
| rs6916779 | Normal | 6 | 32684317 | 6p21.32 | C | T | 8.43E-08 | 8.43E-08 | 0.5888 | 0.5888 | 0.6183 | 0 |
| rs9275618 | Normal | 6 | 32684387 | 6p21.32 | G | A | 2.17E-08 | 1.64E-06 | 0.5455 | 0.5343 | 0.2566 | 26.49 |
| rs9275638 | Normal | 6 | 32684760 | 6p21.32 | T | C | 1.30E-07 | 1.30E-07 | 0.5861 | 0.5861 | 0.5088 | 0 |
| rs3916765 | Normal | 6 | 32685550 | 6p21.32 | A | G | 8.53E-09 | 2.14E-07 | 0.5327 | 0.525 | 0.2939 | 18.33 |
| rs10969985 | Normal | 9 | 30943593 | 9p21.1 | T | C | 6.52E-07 | 0.00471 | 2.1281 | 1.9632 | 0.1071 | 55.25 |
| rs143280565 | Normal | 13 | 63147291 | 13q21.31 | T | G | 1.01E-07 | 1.38E-05 | 2.793 | 2.8335 | 0.2212 | 33.72 |

**Supplementary Table 3 – Risk loci for acute myeloid leukemia (AML) stratified by age.** Table shows fixed effects meta odds ratios for risk loci at 11q13.2 (rs4930561), 1p31.3 (rs10789158), 6p21.32 (rs3916765) and 7q33 (rs17773014) stratified by age. Cases and controls were stratified into those < 55 years and ≥ 55 years. GWAS1 was not included in the meta-analysis for the ≥ 55 age group because the controls were recruited to the 1958 Birth Cohort and were all genotyped at the age of 45 years. For some cases age was not declared or not known. N, number; OR, odds ratio; 95% CI, 95% confidence interval; Q, Cochran’s Q statistic; I^2^, heterogeneity index I^2^; *P*, *P* value.

| **AML risk variant** | **Cytogenetics** | **Age (years)** | **Case, N / Control, N** | **GWAS, N** | **OR (95% CI)** | ***P* value** | **I^2^** | ***P*Q** |
| --- | --- | --- | --- | --- | --- | --- | --- | --- |
| rs4930561 | All AML | All ages | 4018/10488 | 4 | 1.17 (1.11-1.24) | 2.15e-08 | 0.00 | 0.665 |
|  |  | < 55 | 1401/7019 | 4 | 1.21 (1.11-1.32) | 3.0e-05 | 10.07 | 0.34 |
|  |  | ≥ 55 | 1222/3469 | 3 | 1.10 (0.99-1.22) | 0.08 | 0.00 | 0.76 |
|  |  |  |  |  | Test for heterogeneity |  | 46.87 | 0.17 |
| rs10789158 | All AML | All ages | 4018/10488 | 4 | 1.22 (1.13-1.31) | 2.25e-07 | 0.00 | 0.422 |
|  |  | < 55 | 1401/7019 | 4 | 1.22 (1.09-1.38) | 8.8e-04 | 0.00 | 0.82 |
|  |  | ≥ 55 | 1222/3469 | 3 | 1.10 (0.95-1.27) | 0.19 | 0.00 | 0.74 |
|  |  |  |  |  | Test for heterogeneity |  | 16.99 | 0.27 |
| rs3916765 | Normal | All ages | 1287/10488 | 4 | 1.72 (1.46-2.03) | 1.51e-10 | 26 | 0.256 |
|  |  | < 55 | 480/7019 | 4 | 1.64 (1.25-2.16) | 3.5e-04 | 0.00 | 1.00 |
|  |  | ≥ 55 | 518/3470 | 3 | 1.66 (1.27-2.17) | 2.1e-04 | 43.90 | 0.17 |
|  |  |  |  |  | Test for heterogeneity |  | 0.00 | 0.95 |
| rs17773014 | Normal | All ages | 1287/10488 | 4 | 1.26 (1.15-1.37) | 4.09e-07 | 15 | 0.315 |
|  |  | < 55 | 480/7019 | 4 | 1.36 (1.18-1.58) | 4.3e-05 | 50.84 | 0.11 |
|  |  | ≥ 55 | 518/3470 | 3 | 1.18 (1.03-1.37) | 0.02 | 59.09 | 0.09 |
|  |  |  |  |  | Test for heterogeneity |  | 50.10 | 0.16 |

**Supplementary Table 4 – Risk loci for acute myeloid leukemia (AML) stratified by sex.** Table shows fixed effects meta odds ratios for risk loci at 11q13.2 (rs4930561), 1p31.3 (rs10789158), 6p21.32 (rs3916765) and 7q33 (rs17773014) stratified by sex. For some cases sex was not declared or not known. N, number; OR, odds ratio; 95% CI, 95% confidence interval; Q, Cochran’s Q statistic; I^2^, heterogeneity index I^2^; *P*, *P* value.

| **AML risk variant** | **Cytogenetics** | **Sex** | **Case, N / Control, N** | **GWAS, N** | **OR (95% CI)** | ***P* value** | **I^2^** | ***P*Q** |
| --- | --- | --- | --- | --- | --- | --- | --- | --- |
| rs4930561 | All AML | All | 4018/10488 | 4 | 1.17 (1.11-1.24) | 2.15e-08 | 0.00 | 0.665 |
|  |  | Males | 2102/5133 | 4 | 1.18 (1.09-1.27) | 4.5e-05 | 28.77 | 0.24 |
|  |  | Females | 1902/5255 | 4 | 1.17 (1.08-1.27) | 1.1e-04 | 0.00 | 0.67 |
|  |  |  |  |  | Test for heterogeneity |  | 0.00 | 0.88 |
| rs10789158 | All AML | All | 4018/10488 | 4 | 1.22 (1.13-1.31) | 2.25e-07 | 0.00 | 0.422 |
|  |  | Males | 2102/5133 | 4 | 1.27 (1.14-1.41) | 9.8e-06 | 0.00 | 0.83 |
|  |  | Females | 1902/5255 | 4 | 1.18 (1.06-1.31) | 2.4e-03 | 36.69 | 0.19 |
|  |  |  |  |  | Test for heterogeneity |  | 0.00 | 0.34 |
| rs3916765 | Normal | All | 1287/10488 | 4 | 1.72 (1.46-2.03) | 1.51e-10 | 26 | 0.256 |
|  |  | Males | 642/5133 | 4 | 1.45 (1.16-1.80) | 1.1e-03 | 55.10 | 0.08 |
|  |  | Females | 645/5355 | 4 | 2.11 (1.63-2.73) | 1.2e-08 | 0.00 | 0.86 |
|  |  |  |  |  | Test for heterogeneity |  | 78.47 | 0.03 |
| rs17773014 | Normal | All | 1287/10488 | 4 | 1.26 (1.15-1.37) | 4.09e-07 | 15 | 0.315 |
|  |  | Males | 642/5133 | 4 | 1.27 (1.12-1.44) | 2.3e-04 | 0.00 | 0.65 |
|  |  | Females | 645/5355 | 4 | 1.24 (1.09-1.40) | 7.0e-03 | 4.30 | 0.37 |
|  |  |  |  |  | Test for heterogeneity |  | 0.00 | 0.79 |

**Supplementary Table 5 - *cis*-eQTL analysis of rs4930561 in whole blood.** *cis*-eQTL data for loci annotated to within 500kb of rs4930561 derived from whole blood expression data collated by the eQTLGen Consortium (<http://www.eqtlgen.org/cis-eqtls.html>). SNP, single nucleotide polymorphism; QTL, expression quantitative trait loci; Chr, chromosome. ^a^hg19 coordinates; ^b^Unadjusted ­*P* value; ^c^Benjamini-Hochberg corrected *P* value.

| **SNP** | **SNP Position**^a^ | **Assessed allele eQTL** | **Chr** | **Gene** | **Gene Symbol** | **Gene Position^a^** | **P value eQTL^b^** | ***P*_BH_ eQTL^c^** | **Z score** |
| --- | --- | --- | --- | --- | --- | --- | --- | --- | --- |
| rs4930561 | 67931761 | G | 11 | ENSG00000160172 | FAM86C2P | 67565963 | 0.552111 | 0.763212 | 0.5947 |
|  |  |  |  | ENSG00000255306 | RP5-901A4.1 | 67795102 | 1.75E-25 | 4.1E-24 | -10.4334 |
|  |  |  |  | ENSG00000110717 | NDUFS8 | 67801097 | 3.23E-05 | 0.000169 | -4.1564 |
|  |  |  |  | ENSG00000110719 | TCIRG1 | 67812422 | 1.33E-05 | 7.83E-05 | 4.3546 |
|  |  |  |  | ENSG00000255031 | RP11-802E16.3 | 67819718 | 7.19E-16 | 6.76E-15 | -8.0671 |
|  |  |  |  | ENSG00000110721 | CHKA | 67854618 | 2.21E-15 | 1.73E-14 | 7.9291 |
|  |  |  |  | ENSG00000239559 | RPL37P2 | 67450386 | 0.954545 | 0.99533 | 0.0571 |
|  |  |  |  | ENSG00000132746 | ALDH3B2 | 67439152 | 0.432923 | 0.678246 | -0.7841 |
|  |  |  |  | ENSG00000132744 | ACY3 | 67414078 | 0.19532 | 0.437145 | -1.2949 |
|  |  |  |  | ENSG00000167799 | NUDT8 | 67396405 | 0.46968 | 0.700717 | -0.723 |
|  |  |  |  | ENSG00000231793 | DOC2GP | 67381926 | 1.96E-13 | 1.32E-12 | -7.3514 |
|  |  |  |  | ENSG00000110066 | KMT5B | 67951812 | 0.966818 | 0.99533 | 0.0418 |
|  |  |  |  | ENSG00000167792 | NDUFV1 | 67377164 | 0.966339 | 0.99533 | 0.0422 |
|  |  |  |  | ENSG00000084207 | GSTP1 | 67352598 | 0.50605 | 0.720739 | -0.665 |
|  |  |  |  | ENSG00000171067 | C11orf24 | 68034136 | 0.974153 | 0.99533 | 0.0324 |
|  |  |  |  | ENSG00000162337 | LRP5 | 68148410 | 0.853072 | 0.99533 | -0.1852 |
|  |  |  |  | ENSG00000167797 | CDK2AP2 | 67275044 | 0.297134 | 0.537126 | -1.0426 |
|  |  |  |  | ENSG00000110697 | PITPNM1 | 67266486 | 0.028351 | 0.095177 | -2.1923 |
|  |  |  |  | ENSG00000110711 | AIP | 67254543 | 0.25487 | 0.520821 | 1.1386 |
|  |  |  |  | ENSG00000172663 | TMEM134 | 67234283 | 0.227278 | 0.485549 | -1.2073 |
|  |  |  |  | ENSG00000269913 | CTC-1337H24.1 | 67227561 | 0.9469 | 0.99533 | -0.0666 |
|  |  |  |  | ENSG00000175544 | CABP4 | 67223288 | 0.419551 | 0.678246 | -0.8071 |
|  |  |  |  | ENSG00000175514 | GPR152 | 67219486 | 0.841637 | 0.99533 | -0.1997 |
| (Table continued on following page) | | | | | | | | | |
|  |  |  |  | ENSG00000213402 | PTPRCAP | 67204259 | 0.382663 | 0.642327 | 0.873 |
|  |  |  |  | ENSG00000175634 | RPS6KB2 | 67199401 | 0.082 | 0.226705 | -1.7391 |
|  |  |  |  | ENSG00000172531 | PPP1CA | 67177154 | 0.002583 | 0.011038 | 3.0134 |
|  |  |  |  | ENSG00000172508 | CARNS1 | 67187758 | 0.477084 | 0.700717 | -0.711 |
|  |  |  |  | ENSG00000175463 | TBC1D10C | 67174473 | 0.057775 | 0.169715 | -1.8973 |
|  |  |  |  | ENSG00000172613 | RAD9A | 67162528 | 0.613278 | 0.820949 | -0.5052 |
|  |  |  |  | ENSG00000175505 | CLCF1 | 67136643 | 0.757778 | 0.937252 | 0.3084 |
|  |  |  |  | ENSG00000110075 | PPP6R3 | 68305494 | 0.179207 | 0.421137 | 1.3433 |
|  |  |  |  | ENSG00000175482 | POLD4 | 67121345 | 0.282643 | 0.53137 | 1.0745 |
|  |  |  |  | ENSG00000172830 | SSH3 | 67075498 | 0.121911 | 0.301571 | -1.5467 |
|  |  |  |  | ENSG00000172932 | ANKRD13D | 67062987 | 0.027426 | 0.095177 | -2.2053 |
|  |  |  |  | ENSG00000173020 | ADRBK1 | 67043954 | 0.68254 | 0.86701 | 0.4091 |
|  |  |  |  | ENSG00000260808 | CTD-2007L18.5 | 68382273 | 0.893244 | 0.99533 | 0.1342 |
|  |  |  |  | ENSG00000179038 | AP001885.1 | 66964014 | 0.111925 | 0.292249 | 1.5897 |
|  |  |  |  | ENSG00000173120 | KDM2A | 66955940 | 0.365359 | 0.635996 | 0.9053 |
|  |  |  |  | ENSG00000132749 | MTL5 | 68496970 | 0.997128 | 0.997128 | -0.0035 |
|  |  |  |  | ENSG00000110057 | UNC93B1 | 67765513 | 0.000232 | 0.001092 | -3.6809 |
|  |  |  |  | ENSG00000110090 | CPT1A | 68566983 | 0.04056 | 0.127088 | -2.0479 |
|  |  |  |  | ENSG00000006534 | ALDH3B1 | 67786396 | 2.86E-19 | 4.48E-18 | -8.974 |
|  |  |  |  | ENSG00000197345 | MRPL21 | 68665023 | 1.60E-30 | 7.5E-29 | 11.4837 |
|  |  |  |  | ENSG00000132740 | IGHMBP2 | 68689690 | 2.43E-17 | 2.85E-16 | -8.4711 |
|  |  |  |  | ENSG00000162341 | TPCN2 | 68837218 | 0.628812 | 0.820949 | 0.4835 |
|  |  |  |  | ENSG00000259799 | RP11-554A11.9 | 68925299 | 0.002932 | 0.011483 | -2.9747 |

**Supplementary Table 6 – Chromosome 6p21.32 risk locus for acute myeloid leukemia (AML) stratified by *NPM1* and *FLT3* mutation status.** Table shows fixed effects meta odds ratios for the risk locus at 6p21.32 (rs3916765) stratified by NPM1 and FLT3 mutation status. NPM1 and FLT3 mutation status was available for cases recruited to GWAS2 and GWAS4 only. N, number; OR, odds ratio; 95% CI, 95% confidence interval; Q, Cochran’s Q statistic; I^2^, heterogeneity index I^2^.

| **AML risk variant** | **AML cytogenetics** | **Mutation status** | **Case, N / Control, N** | **GWAS, N** | **OR (95% CI)** | ***P* value** | **I^2^** | ***P*Q** |
| --- | --- | --- | --- | --- | --- | --- | --- | --- |
| rs3916765 | All AML | All cases | 4018/10488 | 4 | 1.20 (1.09-1.32) | 1.15e-04 | 37 | 0.187 |
|  |  | NPM1-negative | 422/6205 | 2 | 1.28 (0.97-1.68) | 0.08 | 0.00 | 0.62 |
|  |  | NPM1-positive | 231/6205 | 2 | 1.96 (1.29-2.98) | 1.7e-03 | 1.12 | 0.94 |
|  |  |  |  |  | Test for heterogeneity |  | 63.88 | 0.10 |
|  |  | FLT3-negative | 594/6205 | 2 | 1.26 (1.01-1.58) | 0.04 | 0.00 | 0.85 |
|  |  | FLT3-positive | 271/6205 | 2 | 1.52 (1.07-2.16) | 0.02 | 81.37 | 0.02 |
|  |  |  |  |  | Test for heterogeneity |  | 0.00 | 0.38 |
|  | Normal | All cases | 1287/10488 | 4 | 1.72 (1.46-2.03) | 1.51e-10 | 26 | 0.256 |
|  |  | NPM1-negative | 205/6205 | 2 | 1.32 (0.91-1.92) | 0.14 | 0.00 | 0.63 |
|  |  | NPM1-positive | 206/6205 | 2 | 1.95 (1.26-3.01) | 2.8e-03 | 0.00 | 0.96 |
|  |  |  |  |  | Test for heterogeneity |  | 43.74 | 0.18 |
|  |  | FLT3-negative | 328/6205 | 2 | 1.49 (1.10-2.02) | 0.01 | 0.00 | 0.38 |
|  |  | FLT3-positive | 200/6205 | 2 | 1.84 (1.20-2.81) | 5.1e-03 | 0.00 | 0.52 |
|  |  |  |  |  | Test for heterogeneity |  | 0.00 | 0.43 |

**Supplementary Table 7- *cis*-eQTL analysis of rs10789158 in whole blood.** *cis*-eQTL data for loci annotated to within 500Kb of rs10789158 derived from whole blood expression data collated by the eQTLGen Consortium (<http://www.eqtlgen.org/cis-eqtls.html>). SNP, single nucleotide polymorphism; QTL, expression quantitative trait loci; Chr, chromosome. ^a^hg19 coordinates; ^b^Unadjusted ­*P* value; ^c^Benjamini-Hochberg corrected *P* value.

| **SNP** | **SNP Position^a^** | **Assessed allele eQTL** | **Chr** | **Gene** | **Gene Symbol** | **Gene Position^a^** | **P value eQTL^b^** | **PBH eQTL^c^** | **Z score** |
| --- | --- | --- | --- | --- | --- | --- | --- | --- | --- |
| rs10789158 | 64867602 | C | 1 | ENSG00000162433 | AK4 | 65655530 | 0.0019144 | 0.012561175 | 3.1032 |
|  |  |  |  | ENSG00000226891 | RP11-182I10.3 | 65453033 | 0.04860961 | 0.121411686 | 1.9721 |
|  |  |  |  | ENSG00000088035 | ALG6 | 63868747 | 0.05518713 | 0.121411686 | 1.9175 |
|  |  |  |  | ENSG00000079739 | PGM1 | 64092431 | 0.11328477 | 0.207688745 | 1.5838 |
|  |  |  |  | ENSG00000142856 | ITGB3BP | 63982916 | 0.45349565 | 0.623556519 | 0.7498 |
|  |  |  |  | ENSG00000162434 | JAK1 | 65365549 | 0.67463156 | 0.742094716 | 0.4199 |
|  |  |  |  | ENSG00000185031 | RP11-182I10.4 | 65451030 | 0.76265206 | 0.76265206 | -0.3019 |
|  |  |  |  | ENSG00000158966 | CACHD1 | 65047584 | 0.61468307 | 0.742094716 | -0.5033 |
|  |  |  |  | ENSG00000203965 | EFCAB7 | 64013703 | 0.31163369 | 0.489710084 | -1.0117 |
|  |  |  |  | ENSG00000116675 | DNAJC6 | 65797727 | 0.02633742 | 0.09657054 | -2.221 |
|  |  |  |  | ENSG00000162437 | RAVER2 | 65254846 | 0.00228385 | 0.012561175 | -3.0505 |

**Supplementary Table 8 - *cis*-eQTL analysis of rs17773014 in whole blood.** *cis*-eQTL data for loci annotated to within 500Kb of rs17773014 derived from whole blood expression data collated by the eQTLGen Consortium (<http://www.eqtlgen.org/cis-eqtls.html>). SNP, single nucleotide polymorphism; QTL, expression quantitative trait loci; Chr, chromosome. ^a^hg19 coordinates; ^b^Unadjusted ­*P* value; ^c^Benjamini-Hochberg corrected *P* value.

| **rs17773014** | **134093813** | **G** | **7** | **ENSG00000205060** | **SLC35B4** | **133987943** | **0.02245394** | ***0.06736182*** | **2.2827** |
| --- | --- | --- | --- | --- | --- | --- | --- | --- | --- |
|  |  |  |  | ENSG00000085662 | AKR1B1 | 134135569 | 5.92E-24 | 5.32863E-23 | -10.0931 |
|  |  |  |  | ENSG00000155530 | LRGUK | 133880697 | 0.84461017 | 0.950186441 | -0.196 |
|  |  |  |  | ENSG00000172331 | BPGM | 134348062 | 0.30442046 | 0.524218755 | -1.0269 |
|  |  |  |  | ENSG00000122786 | CALD1 | 134542241 | 0.99888296 | 0.99888296 | -0.0012 |
|  |  |  |  | ENSG00000131558 | EXOC4 | 133344585 | 0.08572368 | 0.19287828 | -1.7184 |
|  |  |  |  | ENSG00000146859 | TMEM140 | 134841737 | 0.34947917 | 0.524218755 | 0.9357 |
|  |  |  |  | ENSG00000122783 | C7orf49 | 134816331 | 0.40937159 | 0.526334901 | -0.825 |
|  |  |  |  | ENSG00000105875 | WDR91 | 134882453 | 0.01182917 | 0.053231265 | -2.517 |
