## Supplementary material for "Genome-wide association study identifies susceptibility loci for acute myeloid leukemia": Lin 2021 Supplementary Figures

Lin et al

### Supplementary Figures

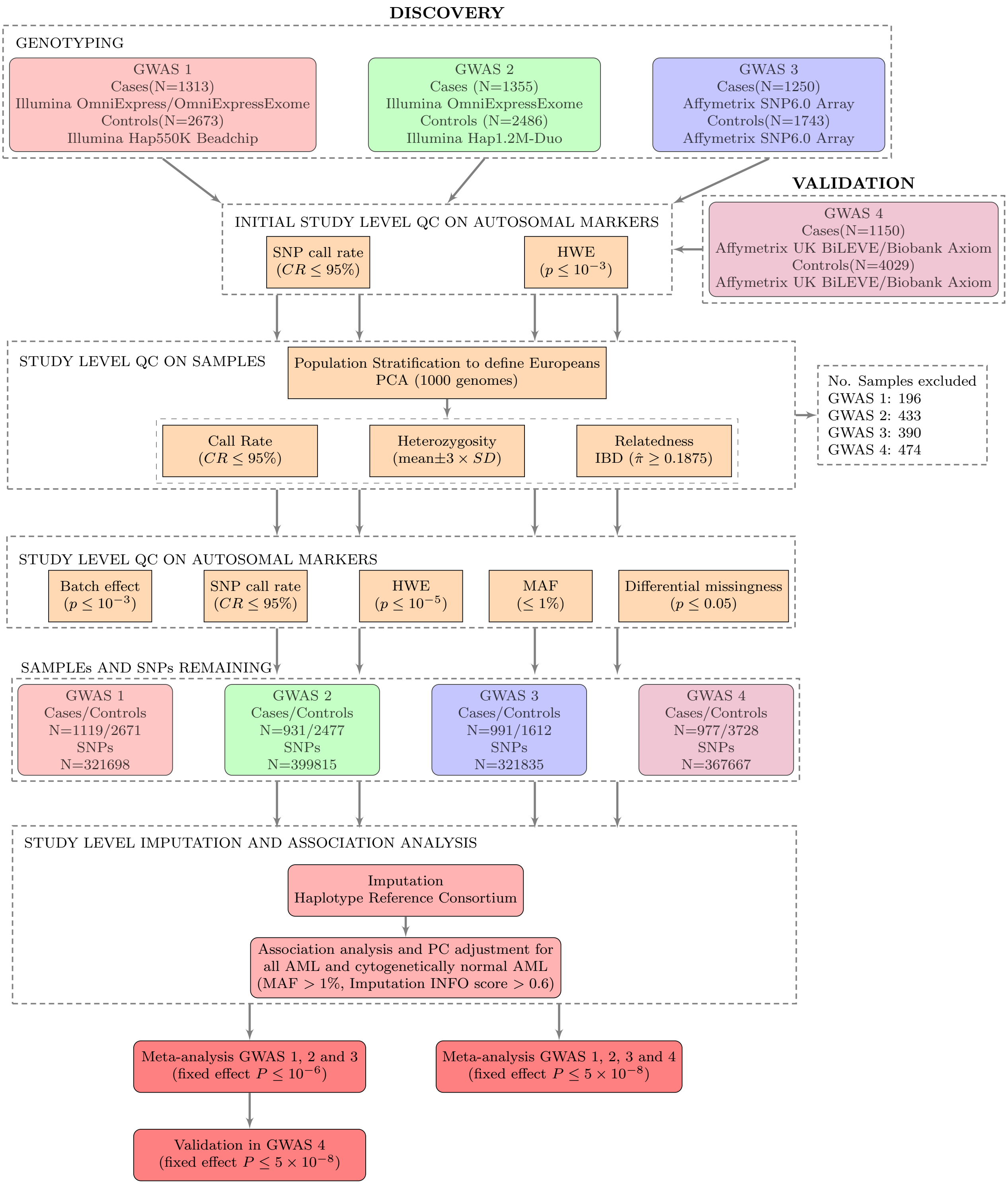


Supplementary Figure 1: **Details of data analysis workflow and quality control filters applied to each AML GWAS.** SNPs with a call rate $< 98\%$ or showing significant deviation from Hardy-Weinberg ($P \leq 10^{-3}$) were excluded. SNPs that showed significant differences ($P < 10^{-3}$) between genotype batches and with significant differences ($P < 0.05$) in missingness between cases and controls were also excluded. Samples were excluded due to low call rate ($< 95\%$), ancestry (principal components analysis), relatedness ($\pi \geq 0.1875$) or heterozygosity (mean $\pm 3 \times$SD). Imputed SNPs with information score $< 0.6$ or MAF $< 0.01$ were excluded.

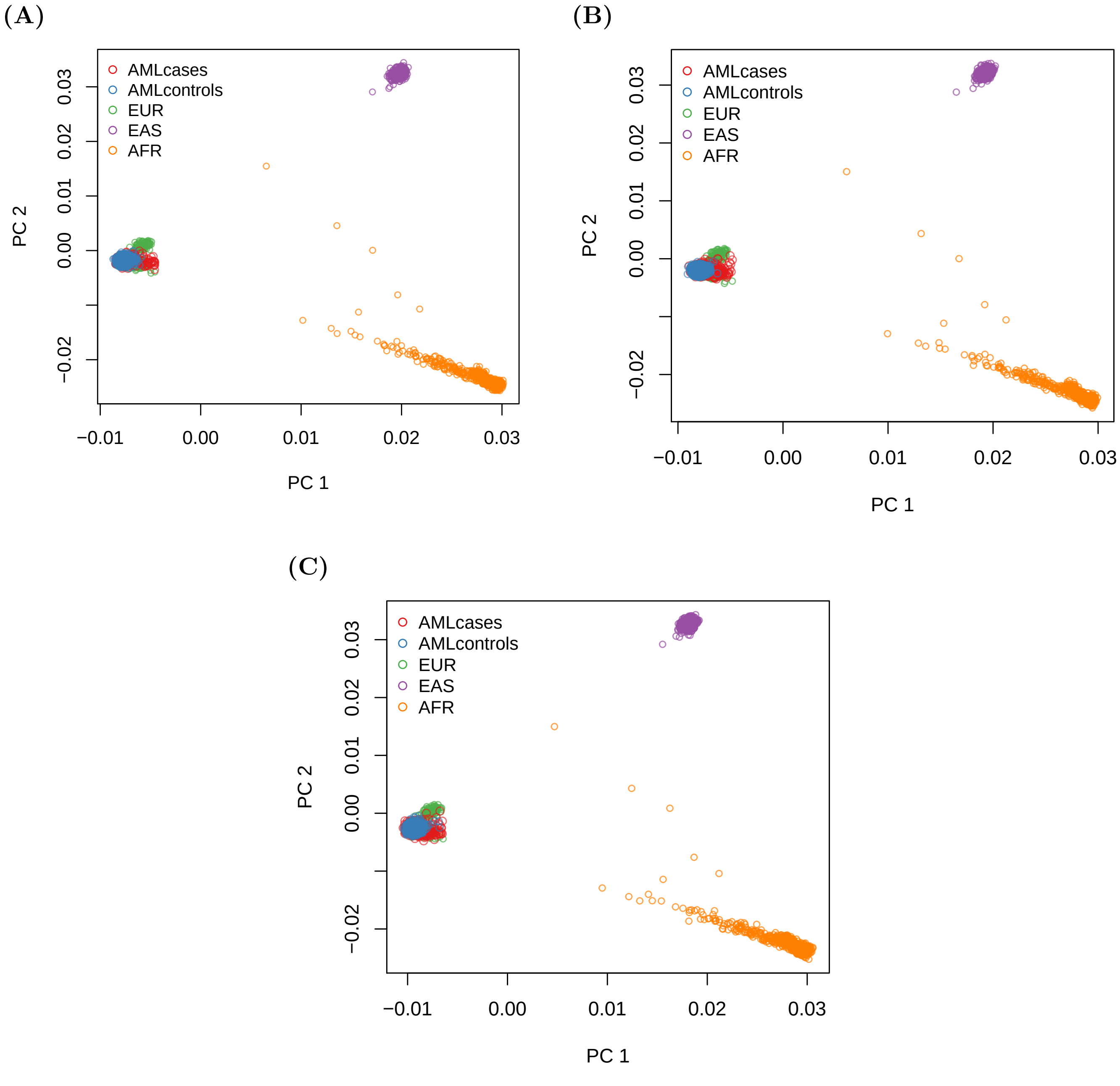


Supplementary Figure 2: **Principal component analysis (PCA) plots of ethnicity structure in (A) GWAS 1 (B) GWAS 2 (C) GWAS 3.** The first two principal components are shown here. European (EUR), East Asian (EAS) and African (AFR) individuals from 1000 genomes project are plotted in green, purple and orange, respectively. AML cases are plotted in red and controls are plotted in blue. PC1, principal component 1; PC2, principal component 2.

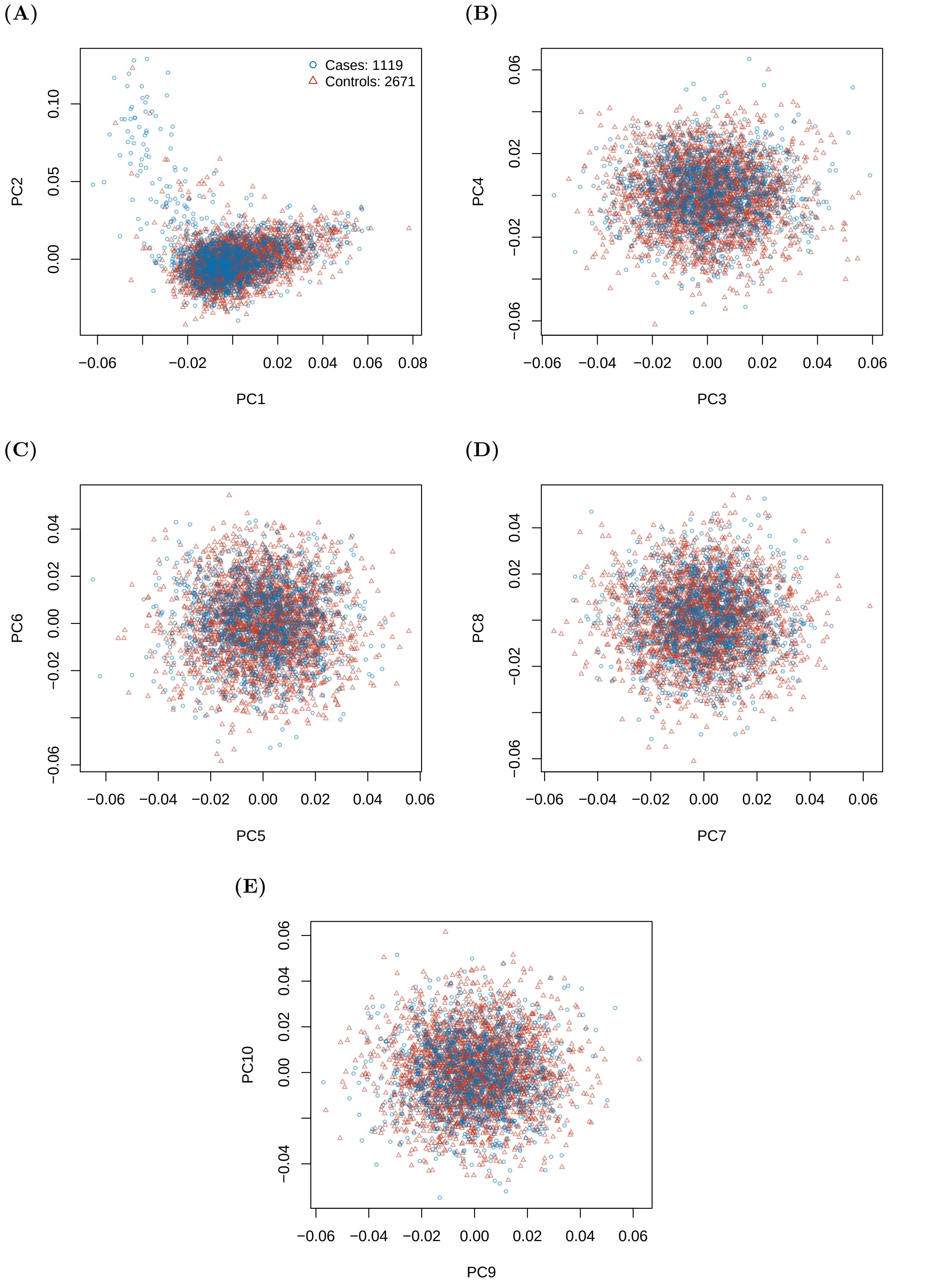


Supplementary Figure 3: PCA in GWAS 1. (A) PC1 and PC2. (B) PC3 and PC4. (C) PC5 and PC6. (D) PC7 and PC8. (E) PC9 and PC10.

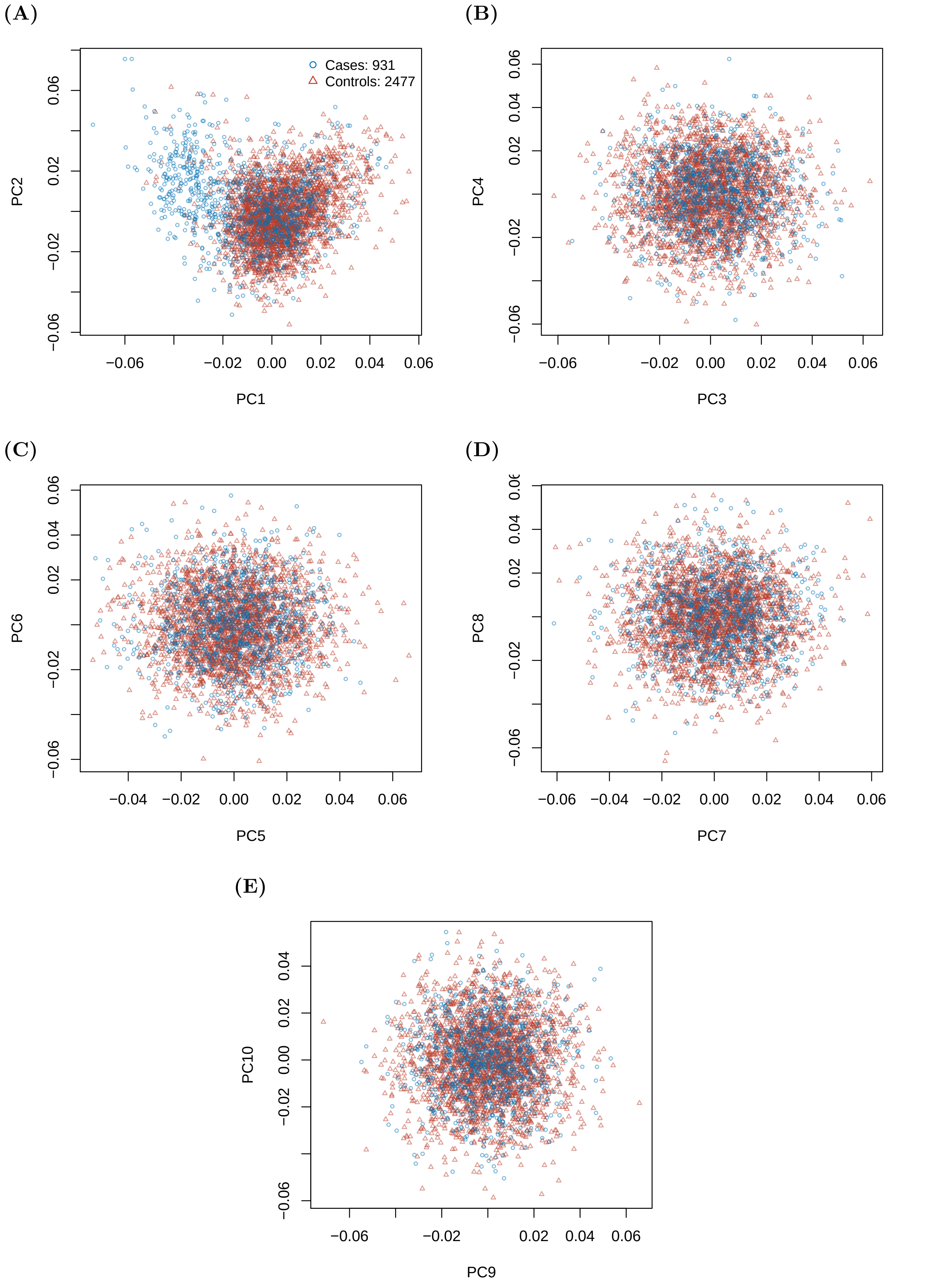


Supplementary Figure 4: PCA in GWAS 2. (A) PC1 and PC2. (B) PC3 and PC4. (C) PC5 and PC6. (D) PC7 and PC8. (E) PC9 and PC10.

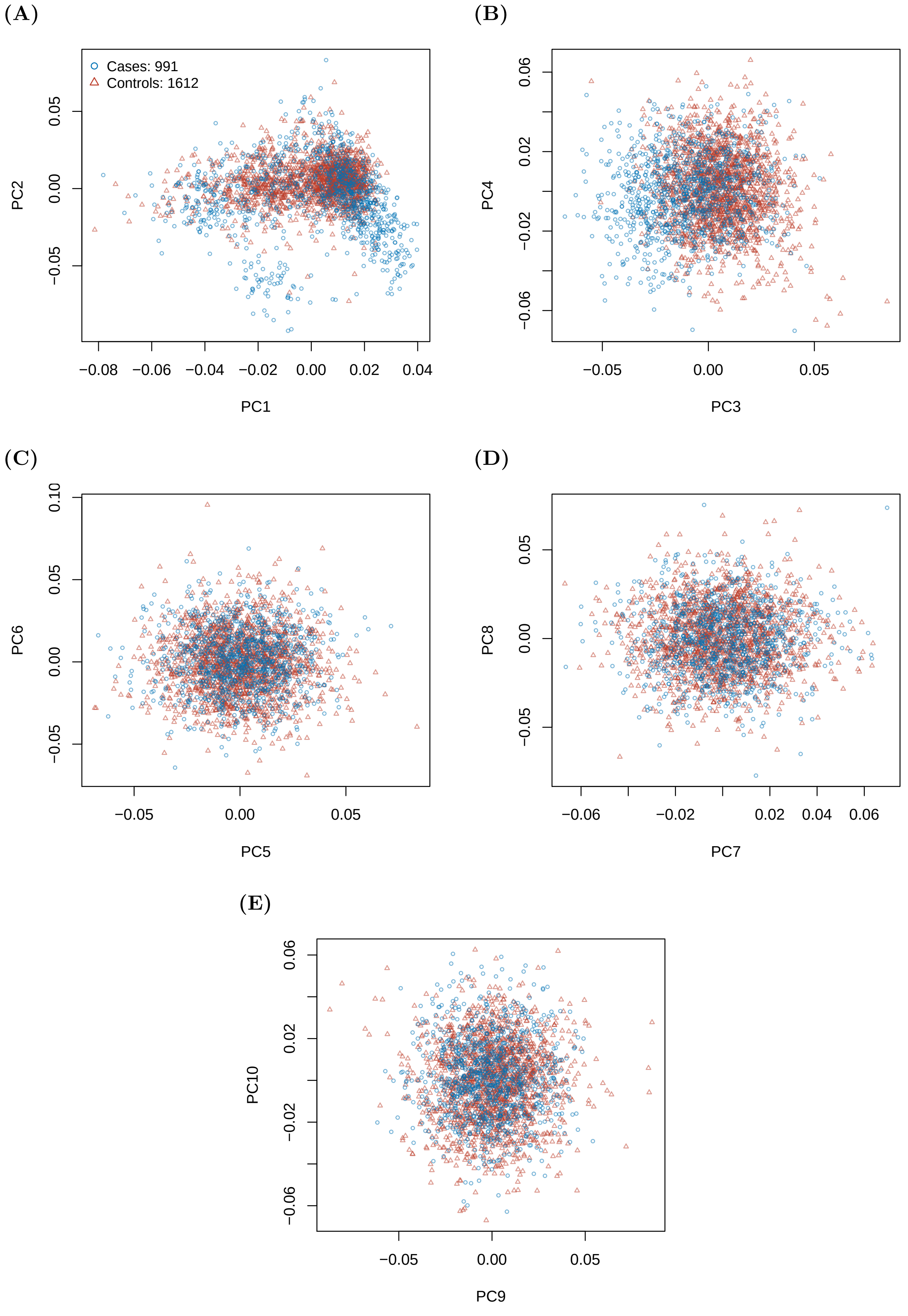


Supplementary Figure 5: PCA in GWAS 3. (A) PC1 and PC2. (B) PC3 and PC4. (C) PC5 and PC6. (D) PC7 and PC8. (E) PC9 and PC10.

(A)

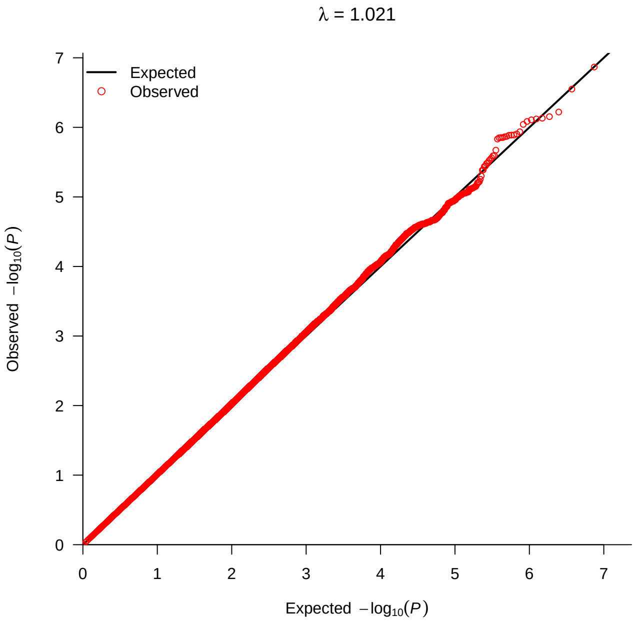


(B)

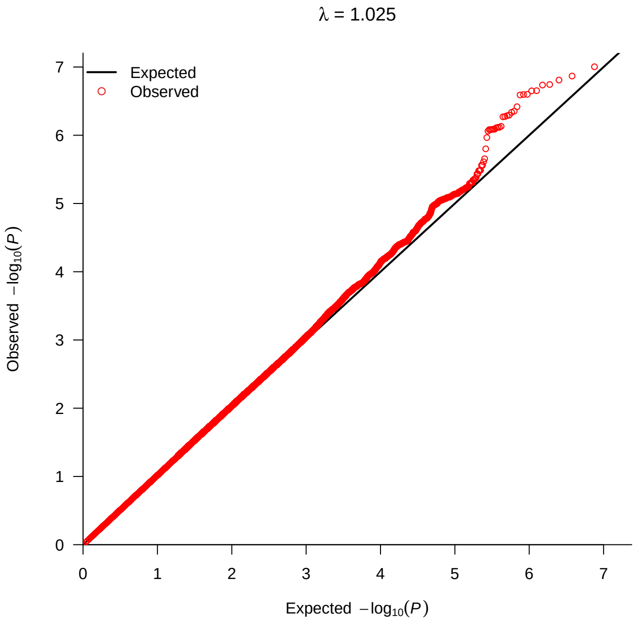


(C)

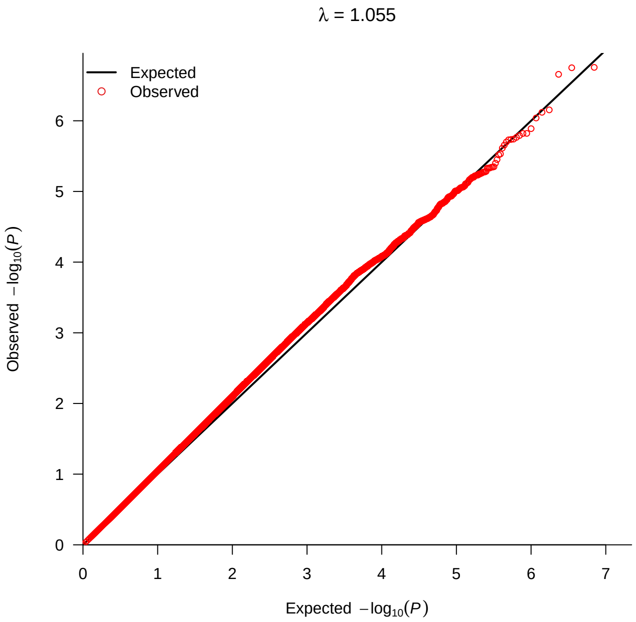


Supplementary Figure 6: **Quantile-Quantile plots of observed P values versus expected P values from association results for all AML in (A) GWAS 1 (B) GWAS 2 (C) GWAS 3.** Association P values (observed versus expected) on imputed genotype data (MAF $> 0.01$, INFO $> 0.6$) are plotted for all AML cases.

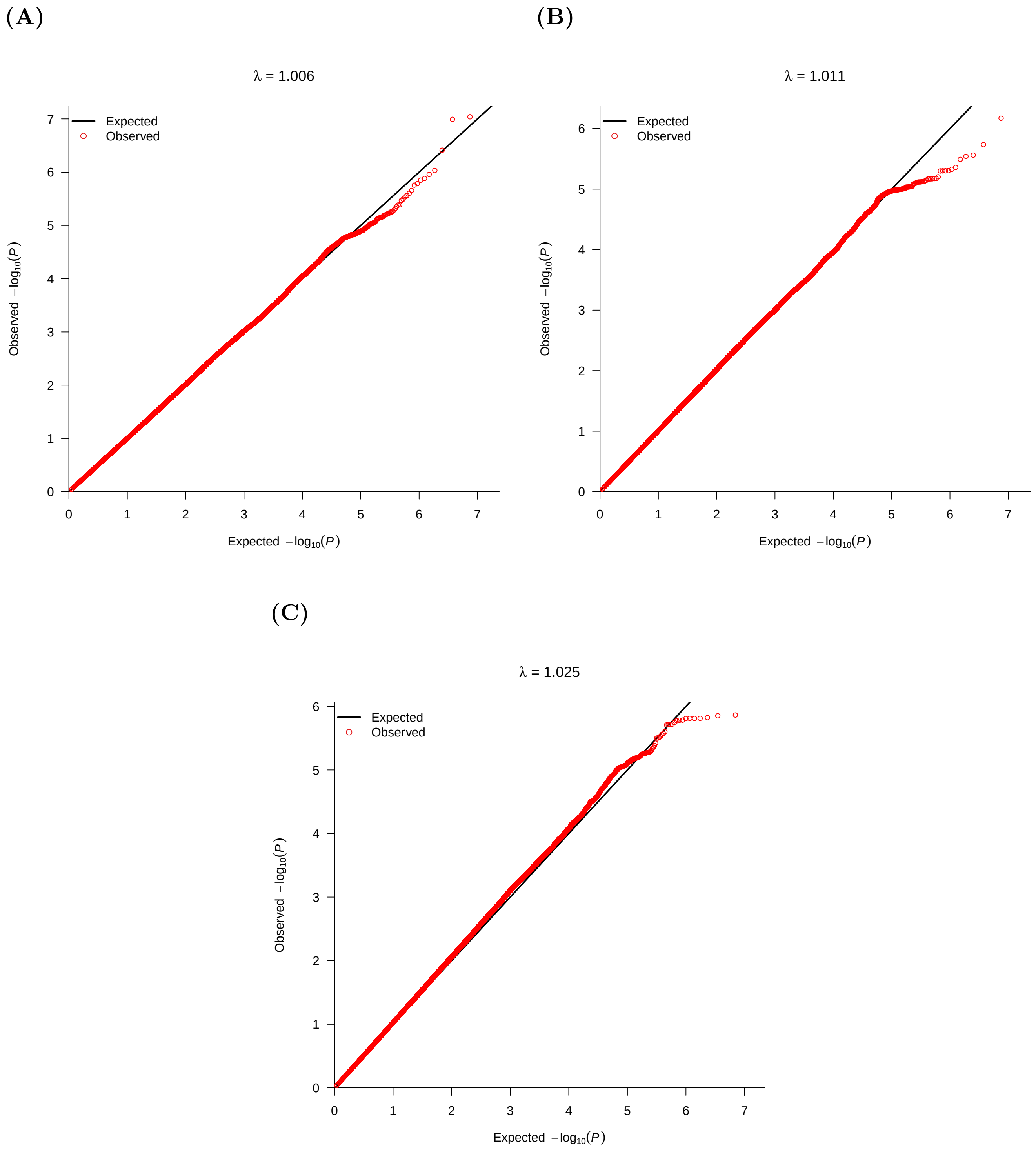


Supplementary Figure 7: **Quantile-Quantile plots of observed P values versus expected P values from association results for cytogenetically normal AML in (A) GWAS 1 (B) GWAS 2 (C) GWAS 3.** Association P values (observed versus expected) on imputed genotype data (MAF $> 0.01$, INFO $> 0.6$) are plotted for cytogenetically normal AML cases.

**(A)**

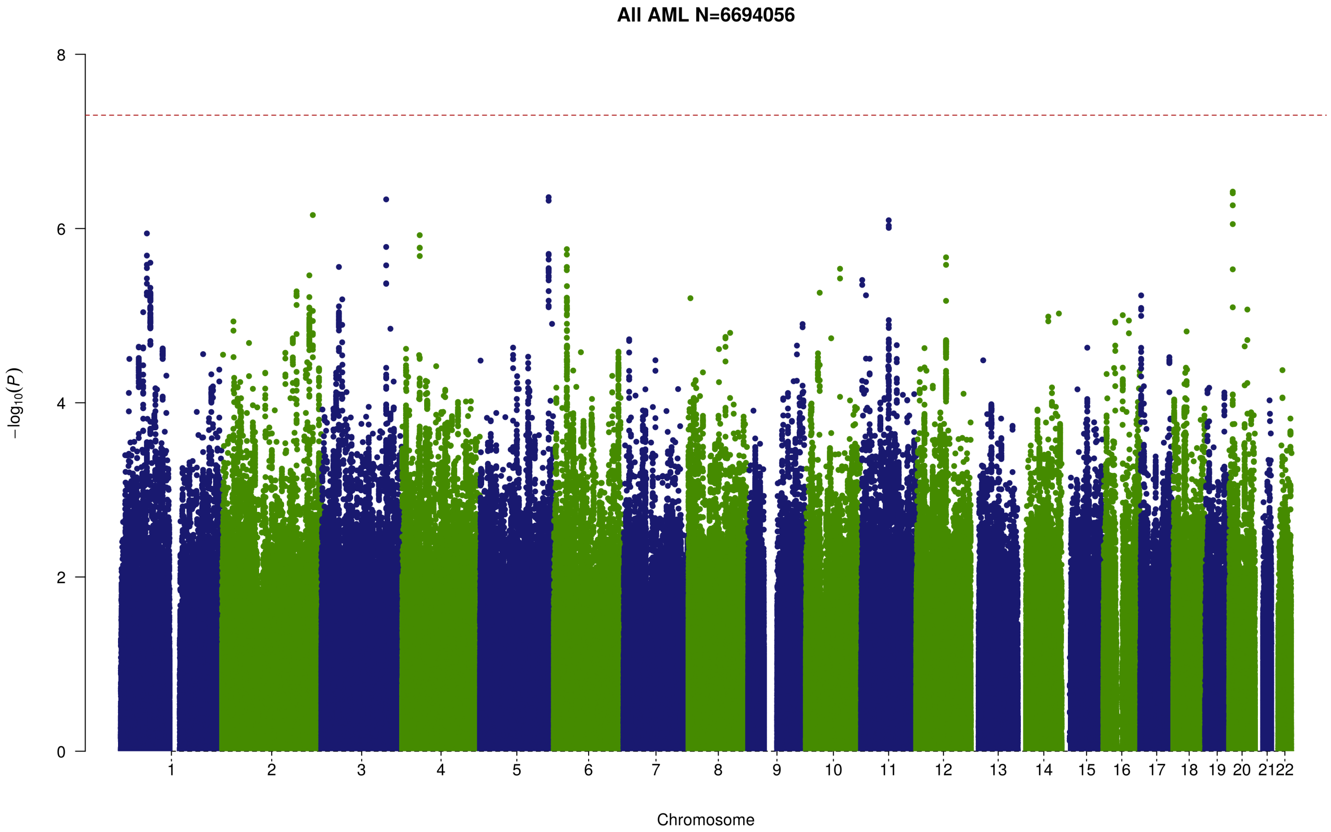


**(B)**

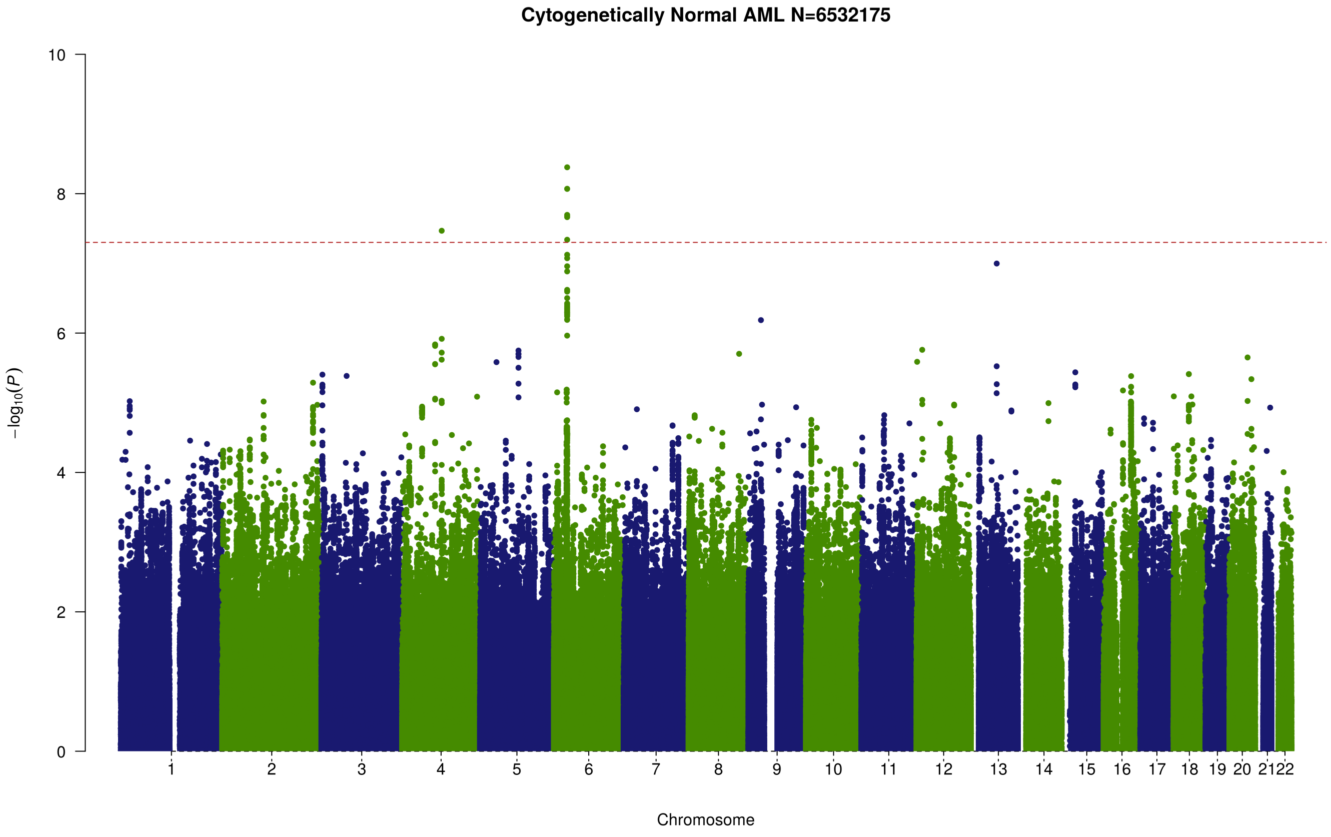


Supplementary Figure 8: **Manhattan plot from meta-analysis of 3 genome-wide association studies for all AML (A) and cytogenetically normal AML (B)**. Manhattan plots show negative $log_{10}$ (fixed effects meta P values, Y-axis) over 22 autosomal chromosomes. Horizontal red line denotes the threshold for statistical significance in a genome-wide association study ($P < 5.0 \times 10^{-8}$).

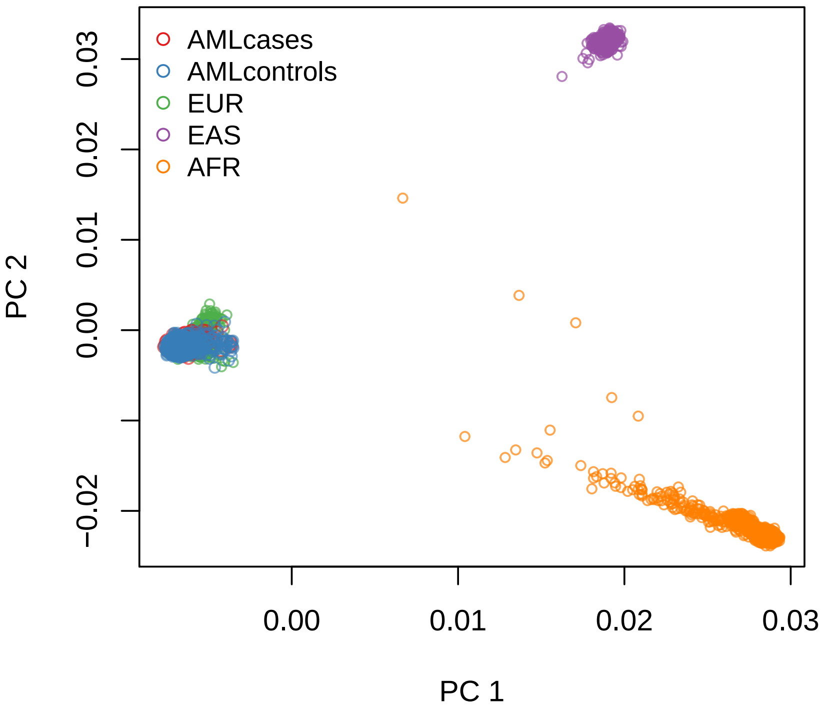


Supplementary Figure 9: **Principal component analysis (PCA) plots of ethnicity structure in GWAS 4.** The first two principal components are shown here. European (EUR), East Asian (EAS) and African (AFR) individuals from 1000 genomes project are plotted in green, purple and orange, respectively. AML cases are plotted in red and controls are plotted in blue. PC1, principal component 1; PC2, principal component 2.

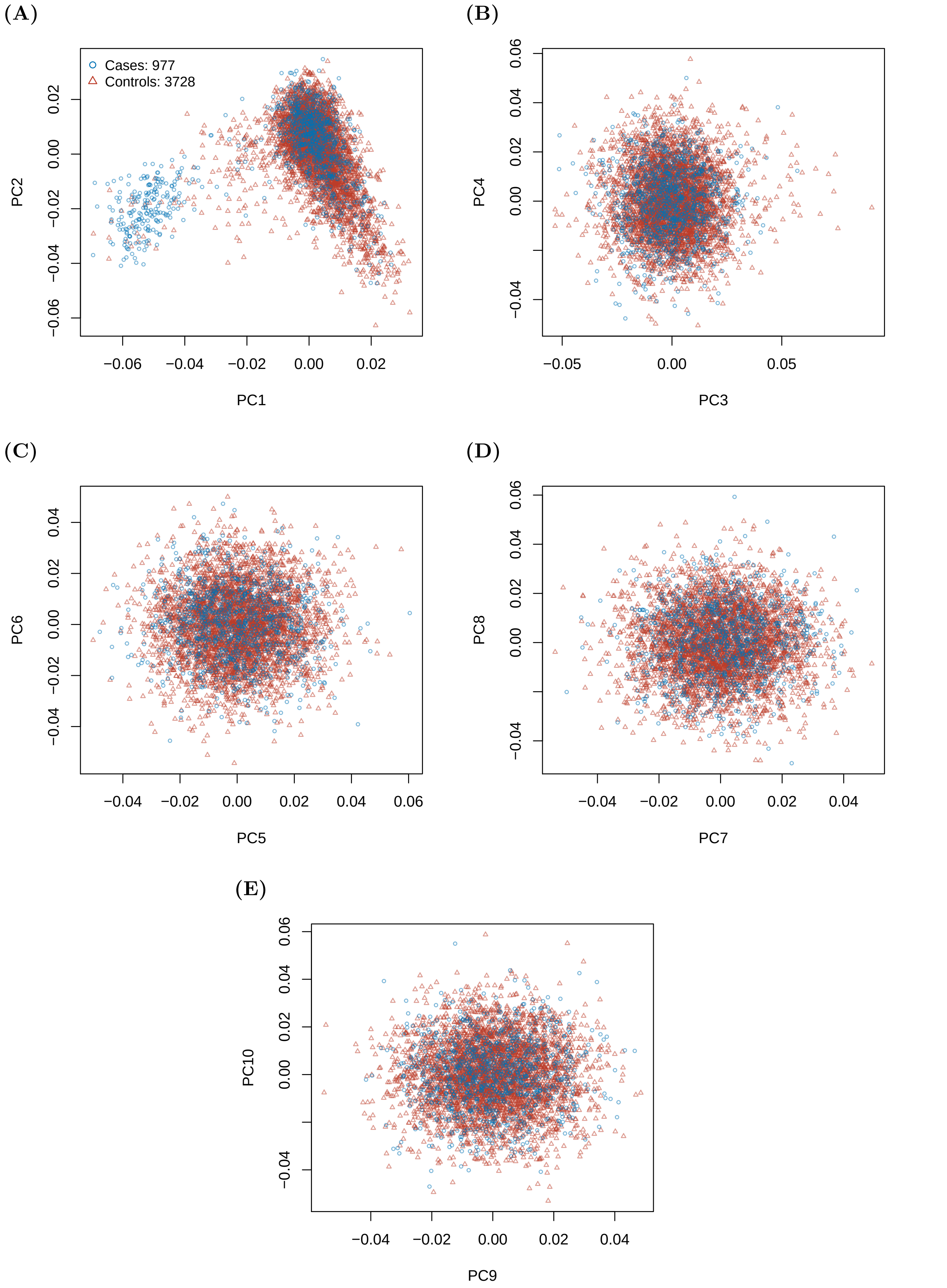


Supplementary Figure 10: PCA in GWAS 4. (A) PC1 and PC2. (B) PC3 and PC4. (C) PC5 and PC6. (D) PC7 and PC8. (E) PC9 and PC10.

**(A)**

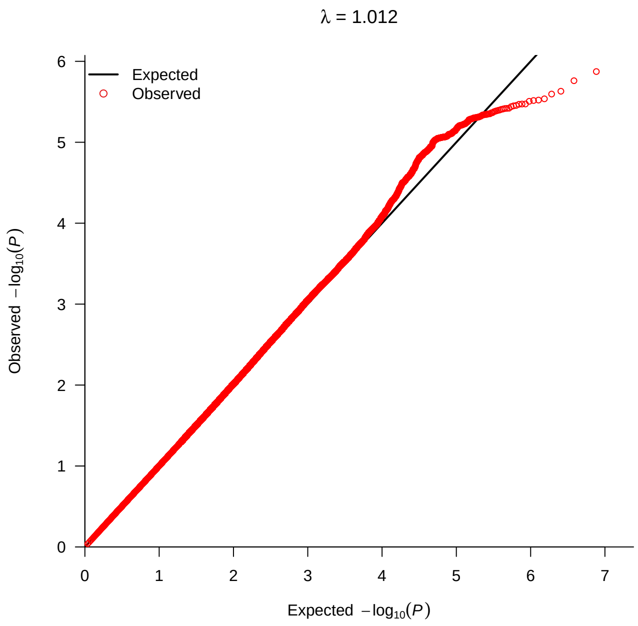


**(B)**

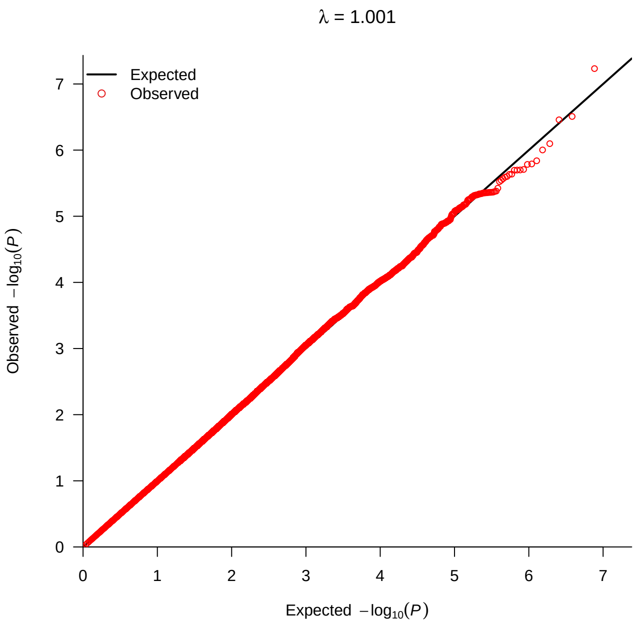


Supplementary Figure 11: **Quantile-Quantile plots of observed P values versus expected P values from association results for all AML (A) and cytogenetically normal AML (B) in GWAS 4.** Association P values (observed versus expected) on imputed genotype data (MAF $> 0.01$, INFO $> 0.6$) are plotted.

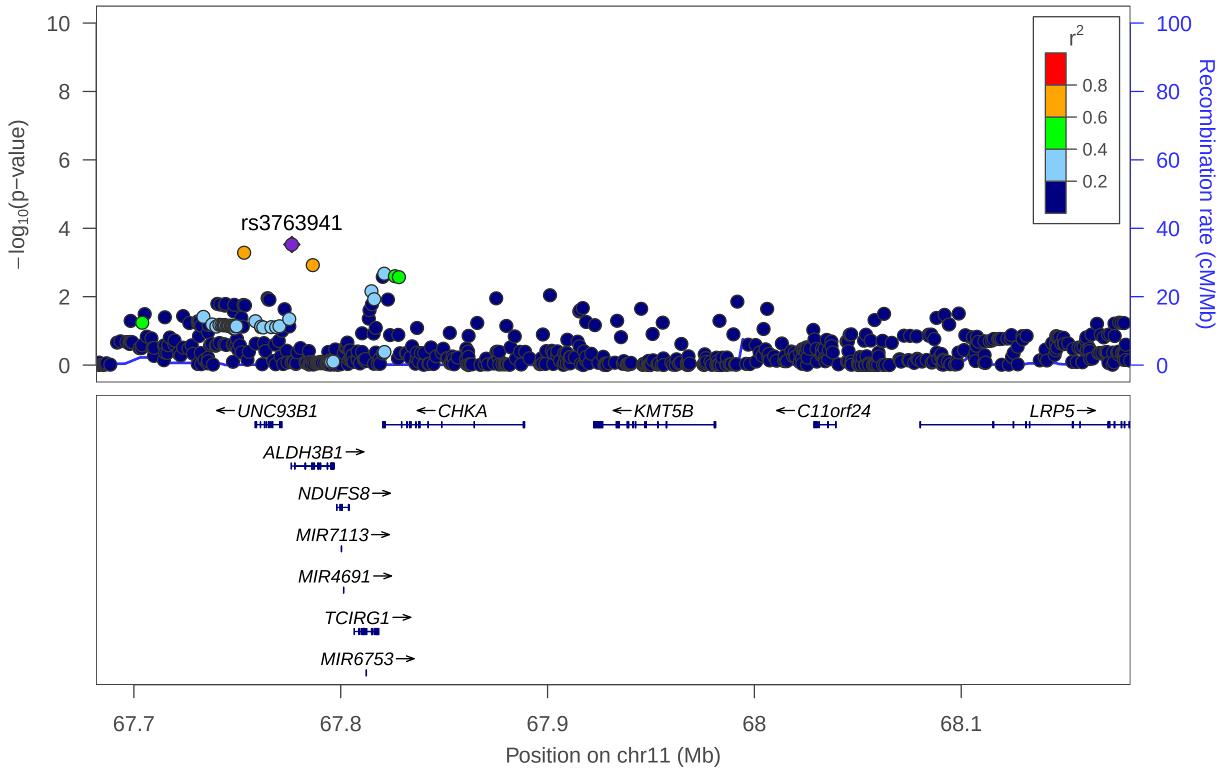


Supplementary Figure 12: **Regional association and linkage disequilibrium plots for association analysis conditioning on the top variant at the chromosome 11q13 susceptibility locus for AML.** Regional association plot showing the chromosome 11q13 AML susceptibility locus conditioning on rs4930561. SNP coordinates based on genomic build b37/h19 are shown on the x-axis and $-log_{10}$ (P values) on the y-axis. SNPs are coloured according to their linkage disequilibrium (pairwise $r^2$) with the lead SNP (annotated) based on the 1000 Genomes European panel. Reference genes in the region are shown in the lower panel, with arrows indicating transcript direction, dense blocks representing exons and horizontal lines representing introns.

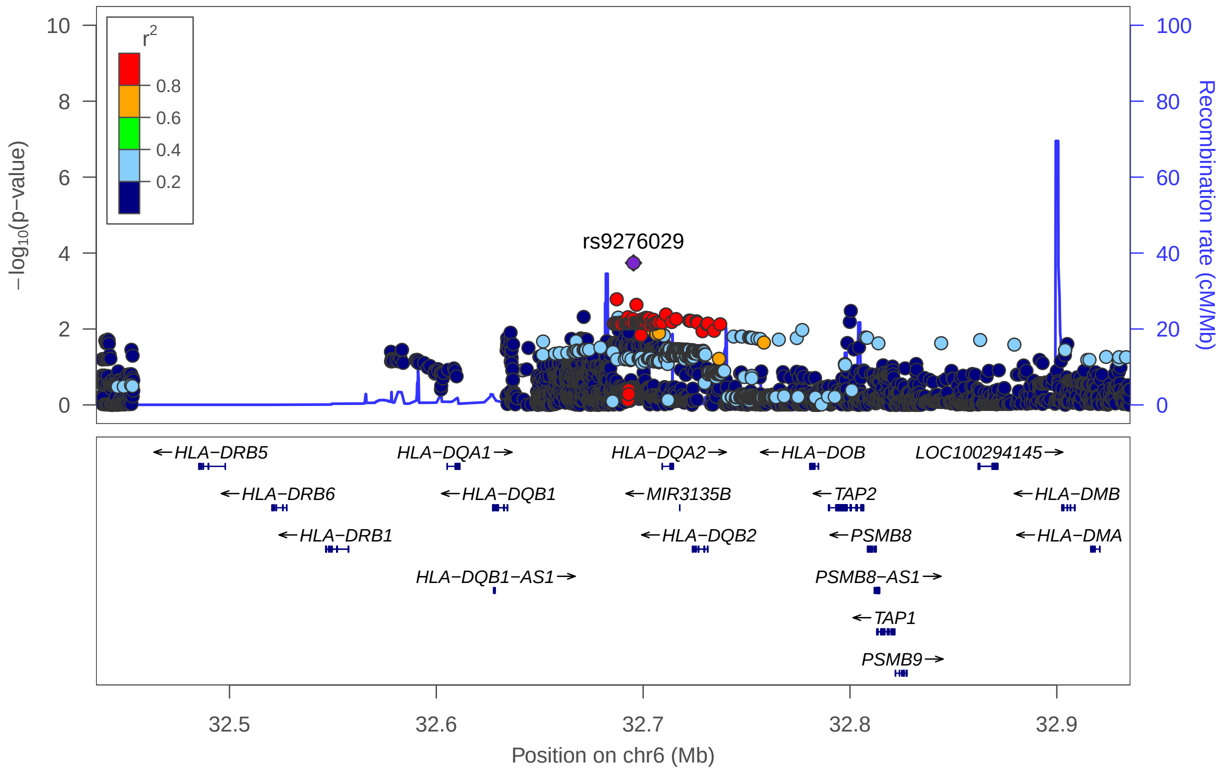


Supplementary Figure 13: **Regional association and linkage disequilibrium plots for association analysis conditioning on the top variant at the chromosome 6p21.32 susceptibility locus for cytogenetically normal AML.** Regional association plot showing the chromosome 6p21.32 AML susceptibility locus conditioning on rs3916765. SNP coordinates based on genomic build b37/h19 are shown on the x-axis and $-log_{10}$ (P values) on the y-axis. SNPs are coloured according to their linkage disequilibrium (pairwise $r^2$) with the lead SNP (annotated) based on the 1000 Genomes European panel. Reference genes in the region are shown in the lower panel, with arrows indicating transcript direction, dense blocks representing exons and horizontal lines representing introns.

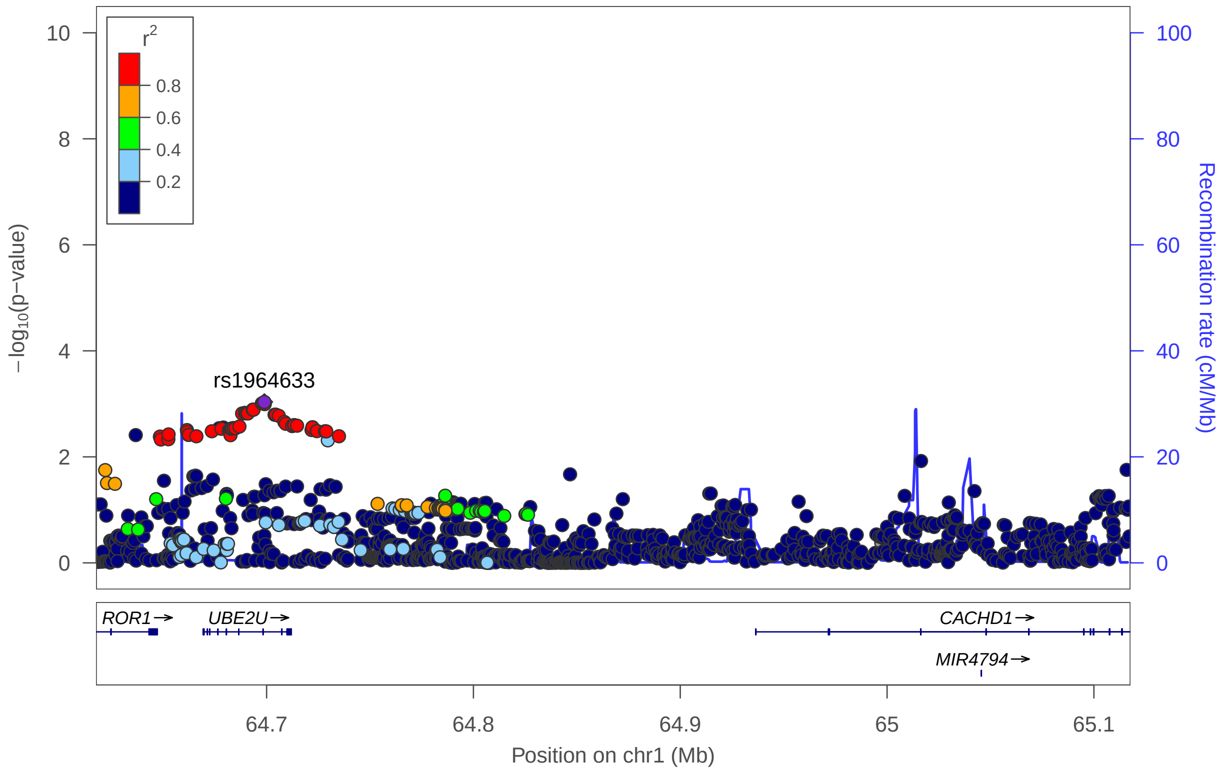


Supplementary Figure 14: **Regional association and linkage disequilibrium plots for association analysis conditioning on the top variant at the chromosome 1p31.3 susceptibility locus for AML.** Regional association plot showing the chromosome 1p31.3 AML susceptibility locus conditioning on rs10789158. SNP coordinates based on genomic build b37/h19 are shown on the x-axis and $-log_{10}$ (P values) on the y-axis. SNPs are coloured according to their linkage disequilibrium (pairwise $r^2$) with the lead SNP (annotated) based on the 1000 Genomes European panel. Reference genes in the region are shown in the lower panel, with arrows indicating transcript direction, dense blocks representing exons and horizontal lines representing introns.

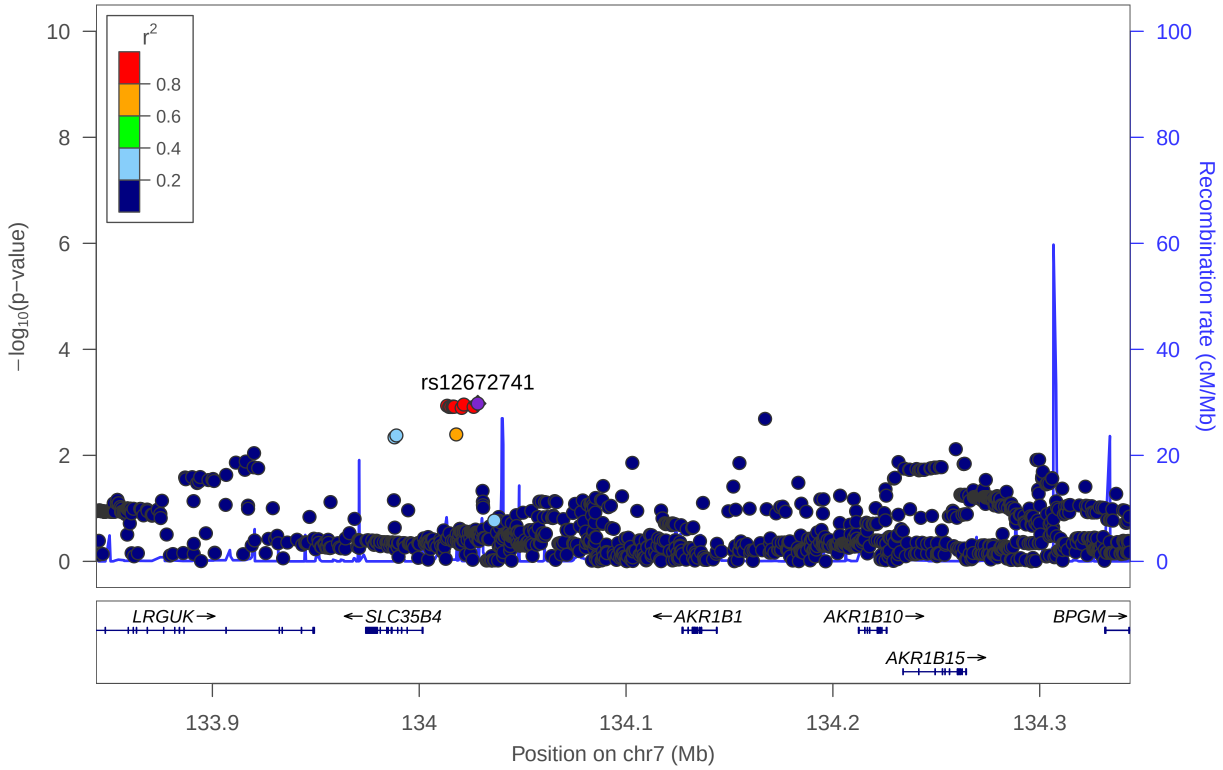


Supplementary Figure 15: **Regional association and linkage disequilibrium plots for association analysis conditioning on the top variant at the chromosome 7q33 susceptibility locus for cytogenetically normal AML.** Regional association plot showing the chromosome 7q33 AML susceptibility locus conditioning on rs17773014. SNP coordinates based on genomic build b37/h19 are shown on the x-axis and $-log_{10}$ (P values) on the y-axis. SNPs are coloured according to their linkage disequilibrium (pairwise $r^2$) with the lead SNP (annotated) based on the 1000 Genomes European panel. Reference genes in the region are shown in the lower panel, with arrows indicating transcript direction, dense blocks representing exons and horizontal lines representing introns.

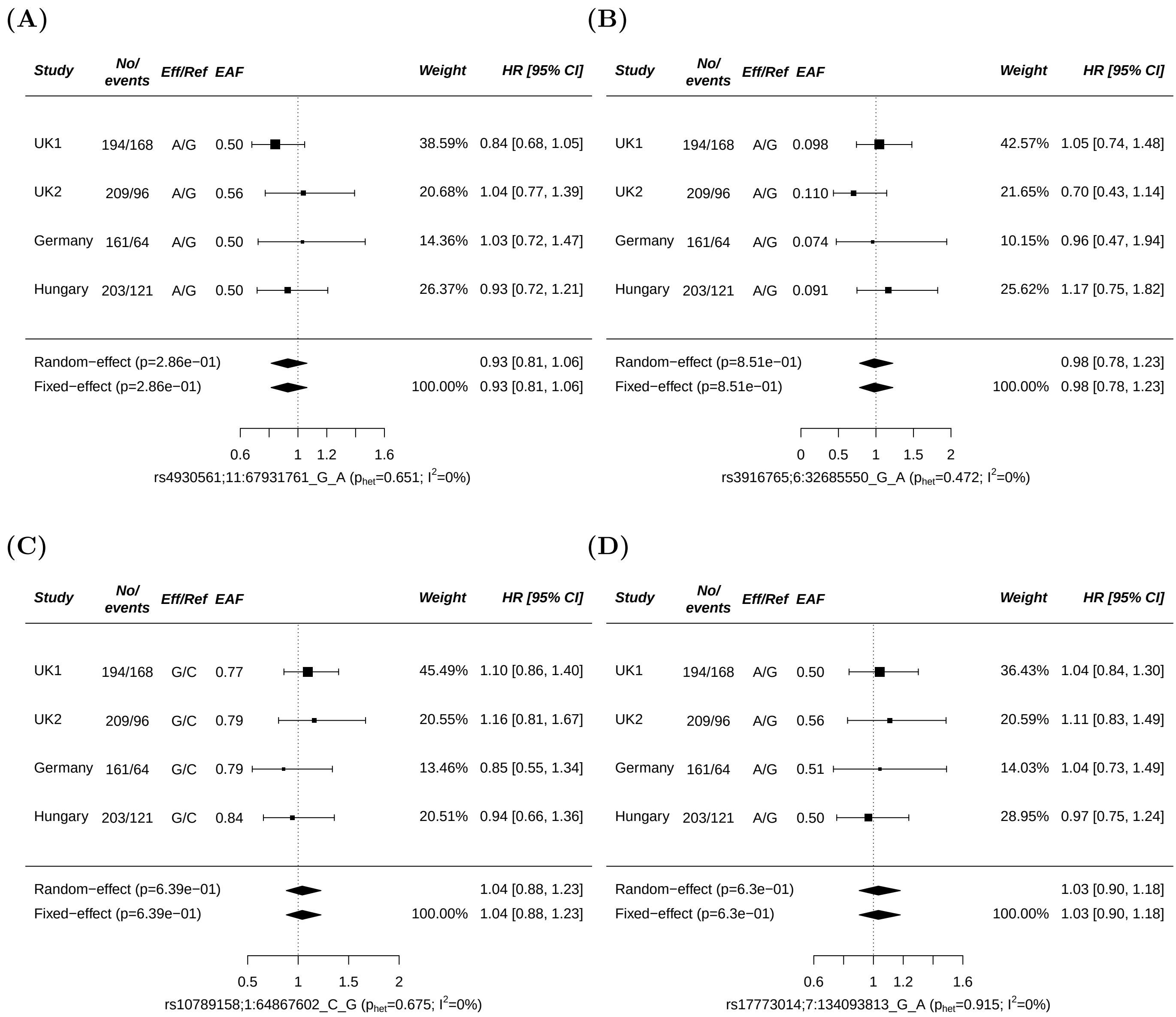


Supplementary Figure 16: **SNP effects on AML overall survival (OS) by study.** (A) rs4930561 (B) rs3916765 (C) rs10789158 (D) rs17773014. Study cohorts (UK1, UK2, Germany and Hungary), number of AML cases (cases), events, effect (Eff) and reference (Ref) allele, effect allele frequencies (EAF) and estimated hazard ratios (HR). The vertical line corresponds to the null hypothesis (HR=1). The horizontal lines and square brackets indicate 95% confidence intervals (95% CI). Areas of the boxes are proportional to the weight of the study. Diamonds represent combined estimates for fixed-effect and random-effect analysis. Cochran's $Q$ statistic was used to test for heterogeneity such that $P_{HET} > 0.05$ indicates the presence of non-significant heterogeneity. The heterogeneity index, $I^2$ (0-100) was also measured which quantifies the proportion of the total variation due to heterogeneity. Overall survival was defined as the time from diagnosis to the date of last follow-up or death (event) from any cause. Cox regression analysis was used to estimate allele specific hazard ratios and 95% CIs.

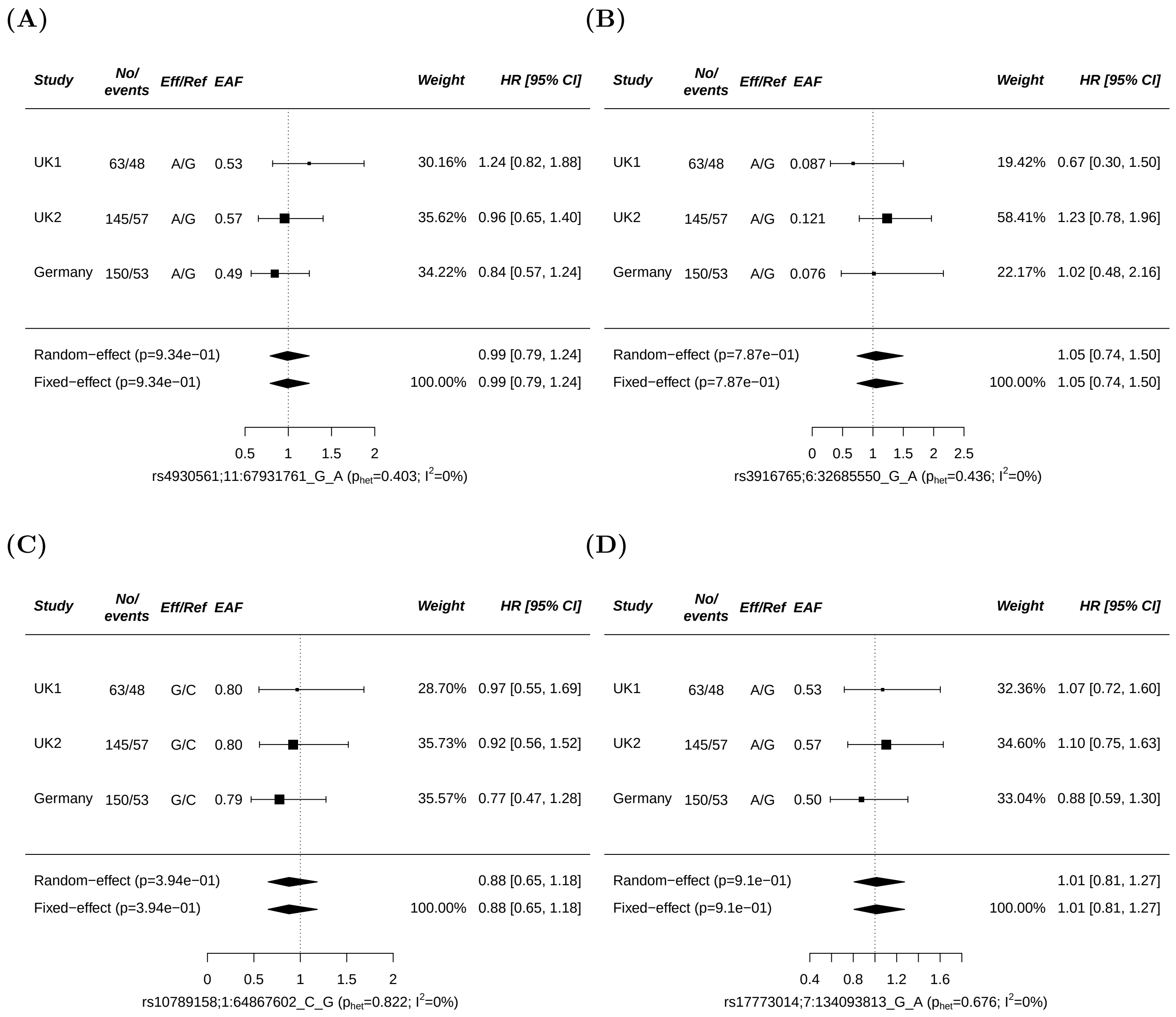


Supplementary Figure 17: **SNP effects on AML relapse-free survival (RFS) by study.** (A) rs4930561 (B) rs3916765 (C) rs10789158 (D) rs17773014. Study cohorts (UK1, UK2, Germany), number of AML cases (cases), events, effect (Eff) and reference (Ref) allele, effect allele frequencies (EAF) and estimated hazard ratios (HR). The vertical line corresponds to the null hypothesis (HR=1). The horizontal lines and square brackets indicate 95% confidence intervals (95% CI). Areas of the boxes are proportional to the weight of the study. Diamonds represent combined estimates for fixed-effect and random-effect analysis. Cochran's $Q$ statistic was used to test for heterogeneity such that $P_{HET} > 0.05$ indicates the presence of non-significant heterogeneity. The heterogeneity index, $I^2$ (0-100) was also measured which quantifies the proportion of the total variation due to heterogeneity. Relapse-free survival was defined as the time from first remission to the date of last follow-up or relapse (event). Cox regression analysis was used to estimate allele specific hazard ratios and 95% CIs.

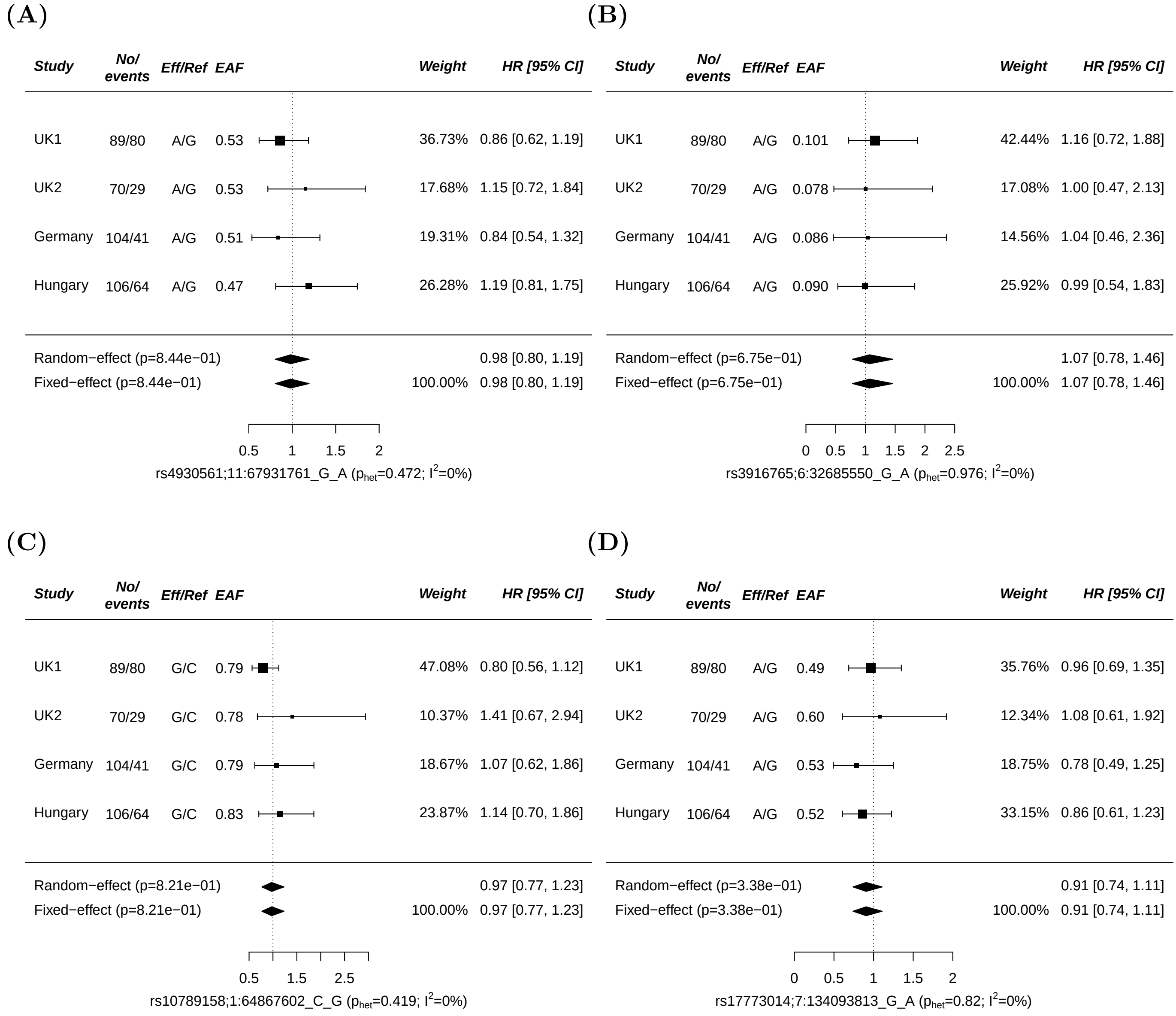


Supplementary Figure 18: **SNP effects on AML overall survival (OS) by study in cytogenetically normal AML.** (A) rs4930561 (B) rs3916765 (C) rs10789158 (D) rs17773014. Study cohorts (UK1, UK2, Germany and Hungary), number of AML cases (cases), events, effect (Eff) and reference (Ref) allele, effect allele frequencies (EAF) and estimated hazard ratios (HR). The vertical line corresponds to the null hypothesis (HR=1). The horizontal lines and square brackets indicate 95% confidence intervals (95% CI). Areas of the boxes are proportional to the weight of the study. Diamonds represent combined estimates for fixed-effect and random-effect analysis. Cochran's $Q$ statistic was used to test for heterogeneity such that $P_{HET} > 0.05$ indicates the presence of non-significant heterogeneity. The heterogeneity index, $I^2$ (0-100) was also measured which quantifies the proportion of the total variation due to heterogeneity. Overall survival was defined as the time from diagnosis to the date of last follow-up or death (event) from any cause. Cox regression analysis was used to estimate allele specific hazard ratios and 95% CIs.

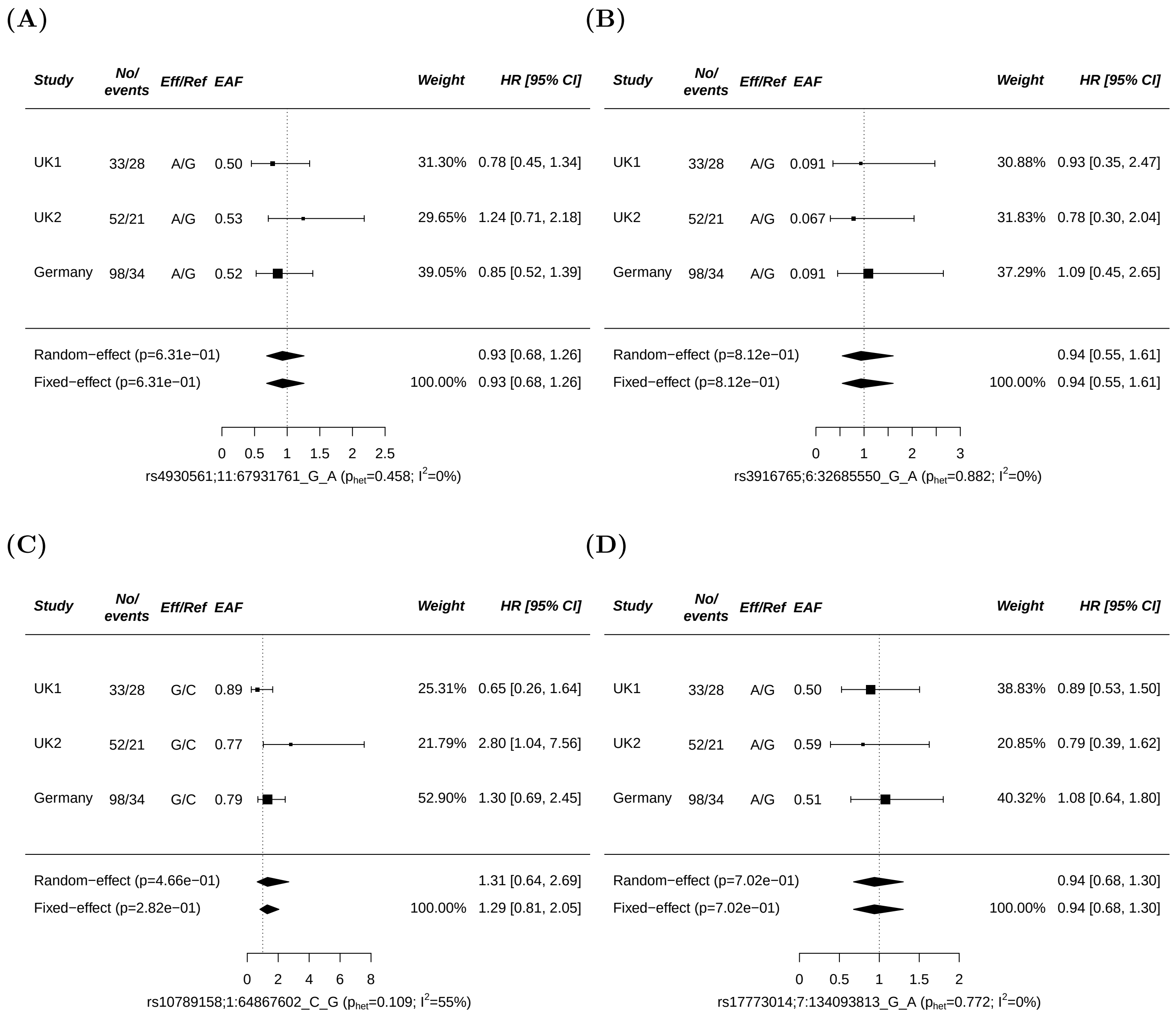


Supplementary Figure 19: **SNP effects on AML relapse-free survival (RFS) by study in cytogenetically normal AML.** (A) rs4930561 (B) rs3916765 (C) rs10789158 (D) rs17773014. Study cohorts (UK1, UK2, Germany), number of AML cases (cases), events, effect (Eff) and reference (Ref) allele, effect allele frequencies (EAF) and estimated hazard ratios (HR). The vertical line corresponds to the null hypothesis (HR=1). The horizontal lines and square brackets indicate 95% confidence intervals (95% CI). Areas of the boxes are proportional to the weight of the study. Diamonds represent combined estimates for fixed-effect and random-effect analysis. Cochran's $Q$ statistic was used to test for heterogeneity such that $P_{HET} > 0.05$ indicates the presence of non-significant heterogeneity. The heterogeneity index, $I^2$ (0-100) was also measured which quantifies the proportion of the total variation due to heterogeneity. Relapse-free survival was defined as the time from first remission to the date of last follow-up or relapse (event). Cox regression analysis was used to estimate allele specific hazard ratios and 95% CIs.

(A)

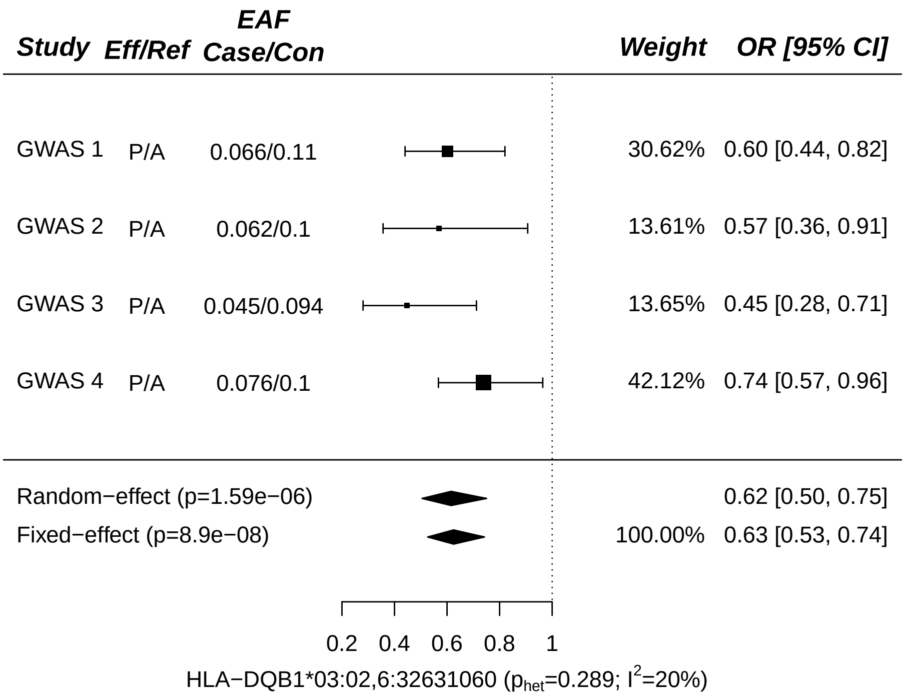


(B)

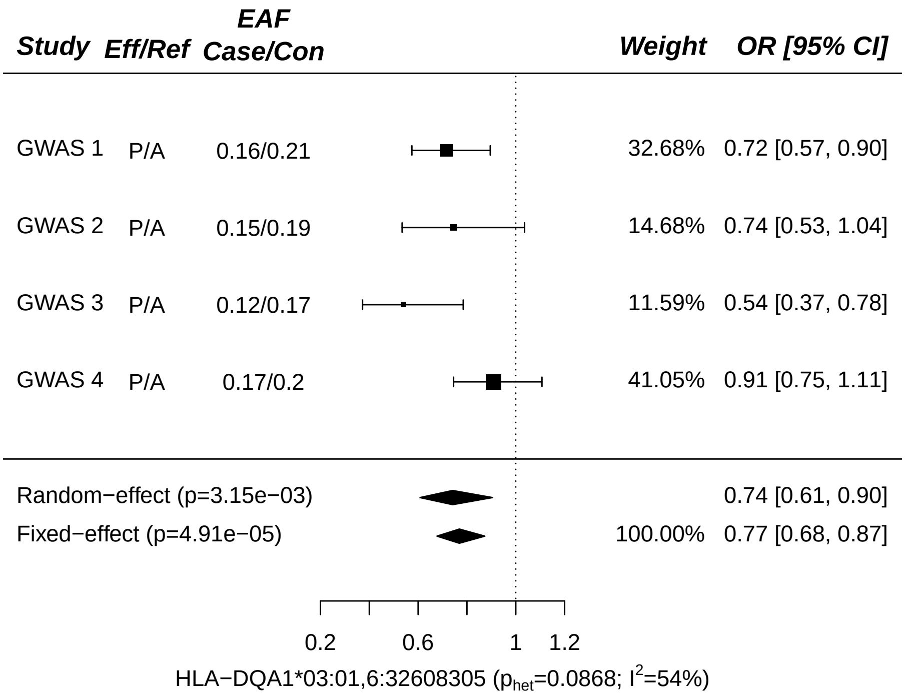


Supplementary Figure 20: **Forest plots for HLA-DQB1*03:02 and HLA-DQA1*03:01 associations with cytogenetically normal AML.** Effect allele frequencies (EAF) and estimated odds ratios (OR) for HLA-DQB1*03:02 (A) and HLA-DQA1*03:01 (B). The vertical line corresponds to the null hypothesis (OR= 1). The horizontal lines and square brackets indicate 95% confidence intervals (95% CI). Areas of the boxes are proportional to the weight of the study. Diamonds represent combined estimates for fixedeffects and randomeffects analysis. Cochran's $Q$ statistic was used to test for heterogeneity such that $P_{HET} > 0.05$ indicates the presence of non-significant heterogeneity. The heterogeneity index $I^2$ (0-100) was also measured which quantifies the proportion of the total variation due to heterogeneity. P, present; A, absent; Eff, effect; Ref, reference.

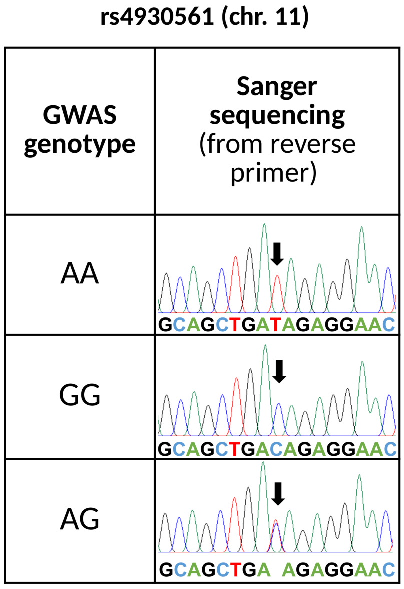


Supplementary Figure 21: **Representative genotype results for rs4930561.** Sanger sequencing was successful for 124 AML cases with 100% (124/124) concordance between GWAS genotyping and Sanger sequencing. Primer sequences for rs4930561 were 5'CCGATTCTTCTGGGGCTTGT3' (forward) and 5'TCTGCAGCATGATTGGAGCA3' (reverse).

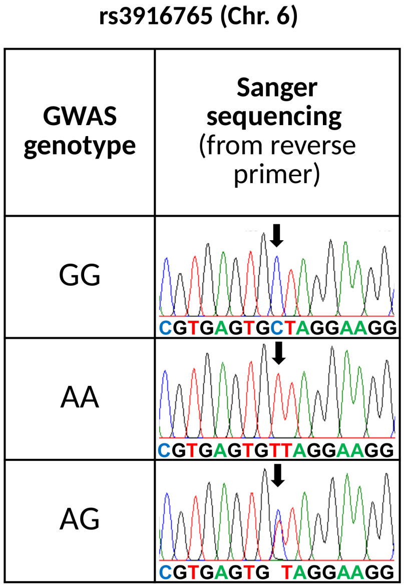


Supplementary Figure 22: **Representative genotype results for rs3916765.** Sanger sequencing was successful for 139 AML cases with 100% (139/139) concordance between GWAS genotyping and Sanger sequencing. Primer sequences for rs3916765 were 5'TTGGTACCTGGGGTATGCTGAA3' (forward) and 5'TGGAGGCTGCCTTGAGATACTA3'(reverse).

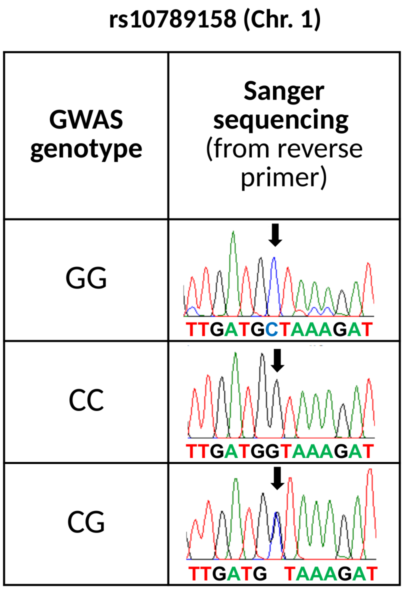


Supplementary Figure 23: **Representative genotype results for rs10789158.** Sanger sequencing was successful for 130 AML cases with 98.5% (128/130) concordance between GWAS genotyping and Sanger sequencing. Primer sequences for rs10789158 were 5'AGCACGTTACAGACTATGCCT3' (forward) and 5'AGCTCAAAGACATGGGGCAA3' (reverse).

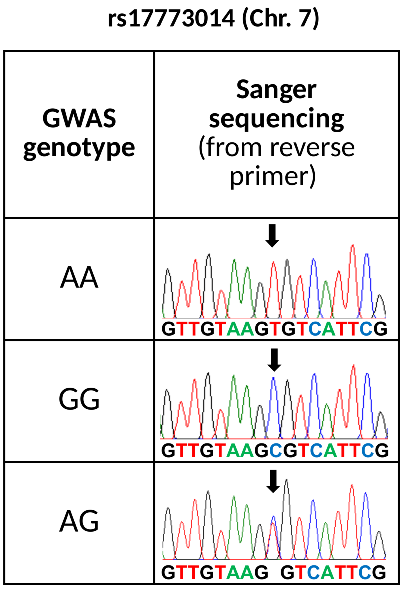


Supplementary Figure 24: **Representative genotype results for rs17773014.** Sanger sequencing was successful for 120 AML cases with 99.2% (119/120) concordance between GWAS genotyping and Sanger sequencing. Primer sequences for rs17773014 were 5'TGTATAACCAAGGGACCGCAC3' (forward) and 5'CACCCCGTCCCATATCCAATG3' (reverse).
